# Brain Cell-Type-Specific X-Chromosome Gene Regulation Reveals Sex-Biased Candidate Genes in Neurodegeneration

**DOI:** 10.64898/2026.11.18.26368437

**Authors:** Youjie Zeng, Noah Cook, Masashi Fujita, Matt Johnson, Aliza P. Wingo, Thomas S. Wingo, Alex R. DeCasien, Cyril Pottier, Carlos Cruchaga, Vilas Menon, Michael E. Belloy

## Abstract

The impact of X-chromosome dosage on gene regulation in specific brain cell-types may importantly shape sex differences in neurodegenerative diseases but remains poorly resolved. We therefore mapped X-chromosome genetic effects on gene expression, i.e. *cis*-expression quantitative trait loci (*cis*-eQTL), across seven brain cell-types from 423 old-age individuals. We identified 212 X-linked genes whose expression was genetically regulated, with the majority being cell-type specific. Notably, 10 of these genes (∼5%) showed sex-heterogeneous genetic regulation. Through sex-differential expression analyses we further discovered 101 sex-biased genes that were predominantly female-upregulated most strikingly in microglia. Sex-biased gene expression showed both shared and distinct cell-type effects, appearing most divergent in non-neuronal populations. Many female-biased genes overlapped known escape-from-X-inactivation genes and were enriched for links to Alzheimer’s disease. Finally, we integrated our resource with X-chromosome-wide association studies of neurodegenerative diseases through causal inference approaches including transcriptome-wide association, mendelian randomization, and colocalization analyses to nominate 11 candidate causal genes, linked to their respective cell-types and sex-informed mechanisms, for Alzheimer’s disease, Parkinson’s disease, and Multiple System Atrophy. Altogether, our findings suggest escape from X chromosome inactivation may contribute to sex differences in neurodegeneration and provide a resource to advance sex-informed insights into disease pathogenesis and therapeutic opportunities.

## INTRODUCTION

Multiple neurodegenerative diseases show prominent sex differences. In terms of prevalence, Alzheimer’s disease (AD) affects more women than men, while inversely Lewy Body dementia (LBD) and Parkinson’s disease (PD) incidence are higher in men^1–3^. Moreover, sex differences are widely reported in clinical presentation, pathology, resilience to pathology, and even treatment response, emphasizing specific genes or mechanisms could contribute to sex-biased disease etiology^4–8^. The X chromosome has been put forward as a factor that may importantly drive such sex differences in brain aging and neurodegeneration^9–11^. This relates in part to females undergoing random X chromosome inactivation (rXCI) of one their X chromosomes to balance gene expression relative to men^12^, but ∼30% of X-linked genes in females display at least some level of escape from inactivation (eXCI)^13–15^. This in turn can lead to double gene dosage and subsequent shifts in biological pathways and phenotypic expression in females^12^. However, due to its complexity and other technical challenges, the X chromosome has historically often been excluded from large-scale genetic association or gene regulation studies^16,17^.

Just in recent years, X-chromosome-wide association studies (XWAS) have begun to successfully implicate X-linked variants in the risk of disorders such as Alzheimer’s disease (AD)^18^, Parkinson’s disease (PD)^19^, Lewy Body dementia (LBD)^20^, and multiple system atrophy (MSA)^21^. To pinpoint candidate causal genes, signals from large-scale genetic association studies are typically integrated with quantitative trait locus (QTL) data mapping genetic variants to gene regulation^22^. While such resources exist for the X chromosome, they remain scarcer than for the autosomes. An important gap remains in mapping X-linked gene regulation in specific brain cell-types. Recent cell-specific QTL studies showed that many genes display cell-type specific regulation which crucially enabled new insights into candidate risk genes and cell-types for neurodegenerative diseases, but the X chromosome was either excluded or not in focus^23,24^. Other cell-type specific studies have started to characterize general sex-biased gene expression patterns and sex-biased gene associations with AD neuropathology and cognitive decline^25^. However, limitations of these efforts variably included small sample sizes, limited exploration of X-linked escape mechanisms, and gaps in determining how sex-biased X-linked genes may relate to neurodegenerative disease risk.

Here, we sought to gain new insights by mapping X-chromosome genetic effects on gene expression, i.e. *cis*-expression quantitative trait loci (*cis*-eQTL), and sex-biased gene expression patterns across seven brain cell-types from old-age individuals (N=423). We specifically assessed evidence for sex-heterogeneous or eXCI effects and related insights from this resource to neurodegenerative disease risk through *in silico* functional genomics and causal inference analyses.

## RESULTS

### A cell-type-specific X chromosome *cis*-eQTL resource in the aged brain

We first sought to map, in a sex-informed manner, how X-linked genetic variation shapes X-linked gene expression in major cell-types in the aged human brain. We leveraged whole genome sequencing (WGS) and matched single-nucleus (sn)RNA-seq data from the dorsolateral prefrontal cortex (DLPFC) of 423 participants in the Religious Orders Study (ROS) and the Memory and Aging Project (MAP) (**Fig.1A**), which included 142 cognitively nonimpaired, 110 mild cognitively impaired, and 171 dementia subjects, with a mean age of 89.2±6.8 years, totaling 288 females and 135 males (cell-type-specific sample sizes and demographics in **Table-S1**). These samples are the same as those used by Fujita *et al*. 2024 to explore genetic regulation of cell-type specific gene expression^24^, but we focused on X-chromosome genetic effects. We conducted non-sex-stratified models that assume rXCI (i.e. male genotype encoding 0/2), sex-stratified analyses, and eXCI modeling within the non-pseudo-autosomal (non-PAR) regions (i.e. male genotype encoding 0/1), while testing the PAR regions 1 and 2 with standard autosomal genotype encoding (**Fig.1A**). Furthermore, we processed the WGS data using X-chromosome-tailored quality-control (QC) and retained variants with minor allele frequency (MAF) ≥1% in all evaluated groups (combined-sex, female, or male), which yielded 224,853 *cis* variants (i.e. those falling within 1Mb of genes for which expression data were available; cf. **Methods**). Showcasing the importance of this QC when using the stricter MAF ≥5% threshold adopted by Fujita *et al*., we still retained 156,968 *cis* variants, a 3.8-fold increase over their 41,234 X chromosome *cis* variants^24^. In contrast, for the autosomes, we would obtain 5,694,179 *cis* variants compared to their 5,562,564 autosomal *cis* variants (98.9% overlap). Across QC and lower MAF thresholds, our X chromosome genotype coverage was therefore 5.5 times larger compared to Fujita *et al*., which is crucial for careful and dense QTL mapping. QTL analyses were then performed using pseudobulked RNA expression, with models adjusted for standard technical and demographic variables, and a two-step false discovery rate (FDR) P<0.05 approach (cf. **Methods**).

**Figure 1.**
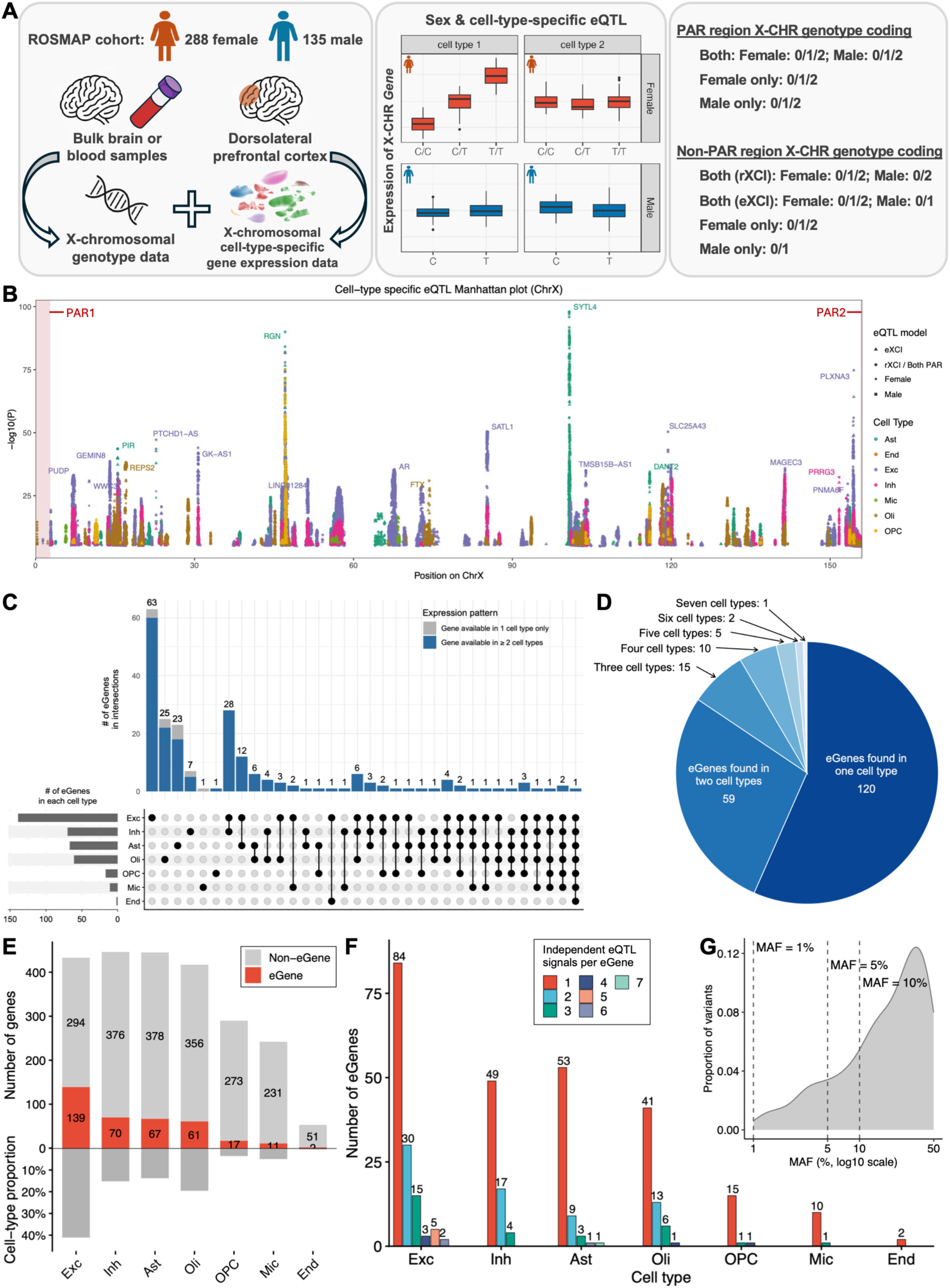
Landscape of X-chromosome-wide cell-type-specific expression quantitative trait loci (eQTLs) in the aged brain. **A)** Schematic of the study design. X-chromosomal genotype data were integrated with cell-type-specific pseudobulk expression data from the dorsolateral prefrontal cortex in the ROSMAP cohort to perform sex-combined/stratified and cell-type-specific eQTL mapping, including assessment of escape from X chromosome inactivation (eXCI). **B)** Manhattan plot showing cell-type-specific eQTL associations across the X chromosome. Only associations passing a two-step FDR threshold are displayed. Points are colored by cell type with marker type indicating the X-chromosome dosage model or sex stratum. The top 20 eGenes (genes with at least one significant eQTL) at respective loci are labeled. **C)** UpSet plot showing eGenes unique to or shared across cell types. Vertical bars represent the number of eGenes in each cell-type intersection. Horizontal bars indicate the total number of eGenes per cell type. Bars are colored by expression availability, distinguishing genes available in one cell type only (gray) from those available in two or more cell types (blue). **D)** Distribution of cell-type-specific eGenes and non-cell-type-specific eGenes. **E)** Top. Number of genes tested per cell type, with genes classified as eGenes or non-eGenes. Bottom. Cell-type proportions. Excitatory neurons showed a higher number of eGenes consistent with their higher cell-type proportion. **F)** Number of eGenes with respective numbers of independent significant eQTL signals in each cell-type. **G)** Minor allele frequency distribution of all independent lead eQTL variants. Dashed lines indicate MAF of 1%, 5%, and 10%.

Within each cell-type, genes having at least one significant *cis*-eQTL were termed eGenes. In total, agnostic to sex or X chromosome inactivation model, we identified 212 unique X-linked eGenes across seven major brain cell-types (**Fig.1B-D**; **Table-S2**), of which 206 were found in the non-PAR region and 6 in the pseudoautosomal regions (5 in PAR1 and 1 in PAR2) (**Fig.1B**, **Table-S3-S5**). Among eGenes detected in only one cell-type, the largest sets were specific to excitatory neurons (63), oligodendrocytes (25), and astrocytes (23) (**Fig.1C**). Importantly, 88.3% (106/120) of cell-type-specific eGenes were expressed in multiple cell-types rather than only in the cell-type where they were detected, confirming the specificity reflects cell-type-specific regulatory activity. In total, of the 212 eGenes, 120 (56.6%) were eGenes in only one of the seven cell-types and 59 in two, whereas just one was shared across all seven (**Fig.1D**). While the proportions of detected eGenes differed across cell-types and appeared highest in excitatory neurons, this pattern was likely in large part due to differing cell-type proportions of assayed nuclei conferring variable power for eGene discovery (**Fig.1E, Table-S2**). Further, across the seven cell-types, we identified 552 independent *cis*-eQTL signals with many eGenes regulated by more than one independent signal (**Fig.1F**). Our observations are consistent with expectations from autosomal studies including the prior work by Fujita *et al*^24^. The maximum of seven independent signals at a single eGene within one cell-type is well within the range seen on the autosomes, where conditional analysis of peripheral blood has resolved up to 13 independent cis-eQTLs at individual genes^26^. Likewise, the finding that eGenes with multiple independent eQTLs were most frequent in the best-powered cell-types matches the tendency for allelic heterogeneity to increase with sample size and statistical power, reaching up to 50% of eGenes in the largest GTEx tissues^27^. Together, these patterns indicate that X-linked *cis*-regulation follows the same power- and frequency-dependent architecture seen for autosomal QTLs.

Finally, to confirm our observations were not driven by inclusion of low-frequency variants, we repeated the analysis under the common-variant threshold of MAF ≥0.05, which reproduced the *cis*-eQTL patterns of the primary analyses (**Fig.S1**). Indeed, most QTL discoveries pertain to variants with MAF ≥ 0.05 (**Fig.1G**). Effect-size estimates were likewise highly concordant between the primary models and models additionally adjusted for cognitive status, indicating minimal influence of the ROSMAP cohort being composed of cognitively normal and cognitively impaired or AD-affected individuals (**Fig.S2**).

### Evidence for sex-biased X-linked *cis*-eQTLs across brain cell-types

We next asked whether X-linked *cis*-eQTLs showed evidence of sex-biased gene regulation or escape from X chromosome inactivation (eXCI). To assess this, we considered which models or sex strata had the highest significance per eQTL and inspected ratios of male-to-female eQTL effect sizes (**Fig.2A-B**). Importantly, during eQTL mapping, male non-PAR genotypes were coded 0/1 and female genotypes 0/1/2. Thus, when assuming rXCI in females, the probability of a genotype being associated with expression and its resultant female effect estimate are inherently reduced by 50%, leading to male-female-beta ratios near 2 (or 1 when log2-transformed). When assuming full eXCI in females, the female genotype association probability is 100% leading to male-female-beta ratios near 1 (or 0 when log2-transformed). Further upward deviations of the male-female-beta ratios then reflect male-biased associations, while further downward ratios reflect female-biased associations. Across all 544 independent non-PAR eQTLs (**Fig.2A, Table-S4**), the majority was most significant in the non-sex-stratified rXCI model (81.1%), but 16.2% were most significant in the eXCI model, 1.7% in females, and 1.1% in males (**Fig.2A**). Notably, these groupings were also largely reflected in their male-to-female effect ratios (**Fig.2A**). The eXCI and female-biased eQTLs pertained to 74 eGenes, of which 37 (50%) overlapped previously reported escape genes (**Table-S6**; cf. **Methods**). However, the remaining 132 eGenes that only related to rXCI or male-biased QTLs had an even larger proportion (N=72, 54.5%) that overlapped previously reported escape genes. While it’s possible that we detected novel escape genes, these findings more likely suggest we were underpowered with the current sample sizes to robustly detect evidence of eXCI through eQTL analyses. Additionally, a gene’s expression from the inactive X chromosome can be relatively modest compared to its expression from the active X chromosome, as escape is quantitative in nature^28,29^, which could further render mapping escape genes through eQTL analyses difficult with limited sample sizes.

**Figure 2.**
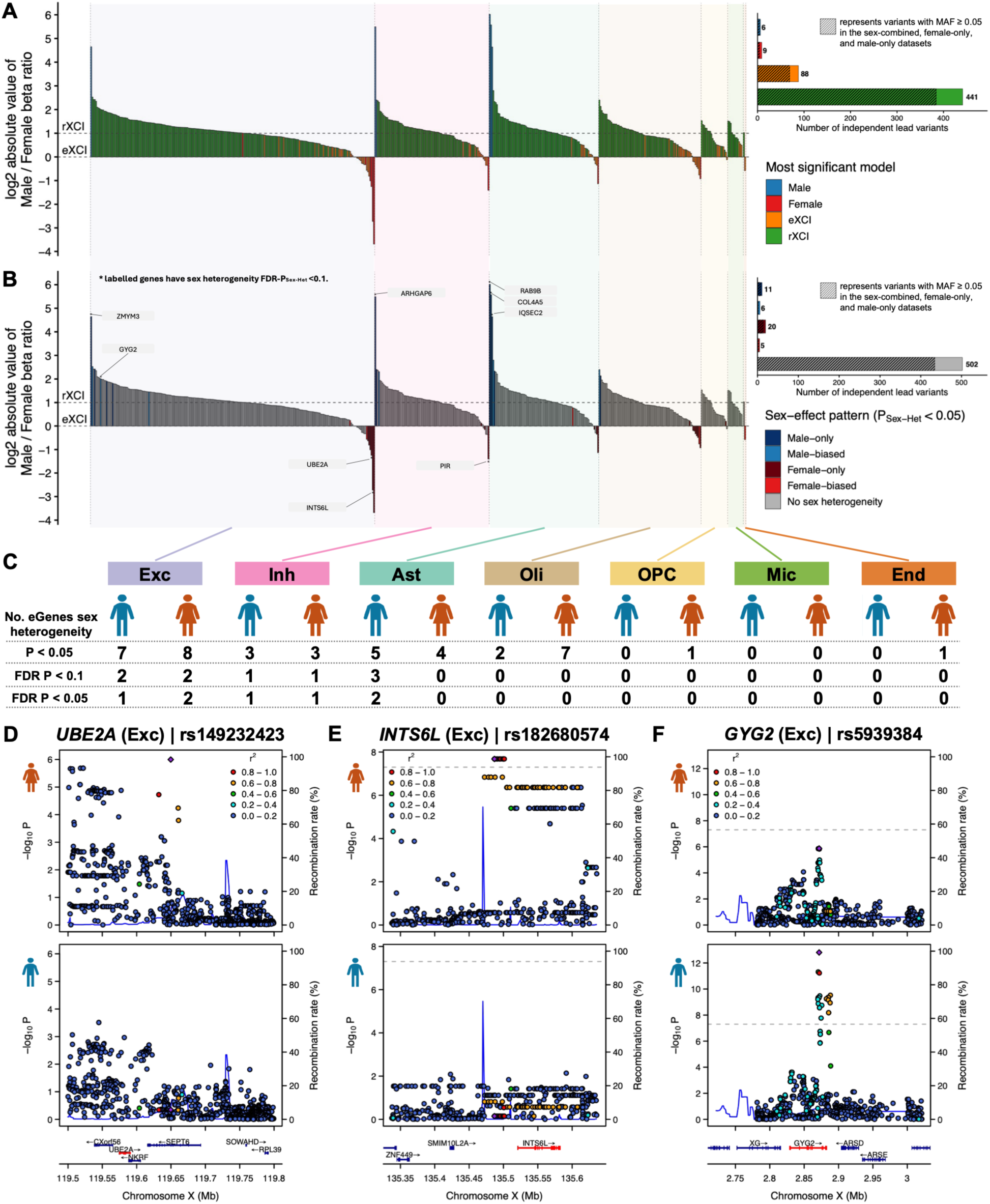
Sex differences in X-linked cell-type specific eQTLs. **A-B)** Left. Across the seven brain cell-types, all independent non-PAR region eQTL signals are shown, ranked within each cell type by the log_2_ absolute ratio of male to female effect size (beta). Log_2_ male-female-beta ratios near 1 are consistent with rXCI assumptions, while higher values reflect male-biased effects; Log_2_ ratios near 0 are consistent with eXCI assumptions, while smaller values reflect female-biased effects (cf. Methods). Right. Dashed boxes indicate variants with MAF≥5%. **A)** Each eQTL is colored by its most significant association model (female, male, eXCI, or rXCI). Bar plots show the distribution of the most significant model across signals. **B)** Same data as in (A) but highlighting signals that pass sex-heterogeneity testing at P_Sex-Het_<0.05 (cf. legend); eQTLs additionally passing FDR-P_Sex-Het_ <0.1 are marked with an asterisk and labeled with the corresponding eGene. Bar plots show the number of independent signals per sex-heterogeneity category. **C)** Number of eGenes with male- or female-biased sex heterogeneity at P_Sex-Het_<0.05, FDR-P_Sex-Het_<0.1, and FDR-P_Sex-Het_ <0.05 in each cell-type. **D-F)** Locus zoom plots (top, female; bottom, male) for three example eGenes with significant sex heterogeneity (FDR-P_Sex-Het_<0.1) in excitatory neurons: **D)** *UBE2A,* **E)** *INTS6L*, and **F)** *GYG2*. *Abbreviations: Exc, excitatory neurons; Inh, inhibitory neurons; Ast, astrocytes; Oli, oligodendrocytes; OPC, oligodendrocyte progenitor cell; Mic, microglia; End, endothelial cells; MAF*, minor allele frequency.

We next sought to identify sex-biased X-linked eQTLs that displayed more extreme male-to-female beta ratios and significance on sex heterogeneity testing (P_Sex-Het_). For the sex heterogeneity tests, we compared male and female effect estimates on the rXCI scale (cf. **Methods**). For the non-PAR regions, at nominal heterogeneity significance (P_Sex-Het_<0.05), 42 eQTLs appeared sex-biased with roughly equal male and female-biased effects (**Fig.2B-C**). After further multiple comparison correction, 9 eQTLs or eGenes appeared sex-biased at FDR-P_Sex-Het_<0.1 and 7 eQTLs or eGenes were sex-biased at FDR-P_Sex-Het_<0.05. Example locus zoom plots for 3 sex-biased eQTLs are shown in **Fig.2D-F**. The 9 eGenes observed with FDR-P_Sex-Het_<0.1 pertained to 5 male-only, 1 male-biased (*GYG2*), and 3 female-only eQTLs and were found in neurons and astrocytes, consistent with those cell-types having increased power for eGene discovery (**Fig.1E**). The 3 female-only eQTLs showed effect size ratios at least 4 times larger or in opposite direction relative to males, supporting they reflected truly female-biased association patterns rather than just different dosage models across males and females. Further, 7 out of these 9 sex-biased eGenes were previously reported escape genes, while additionally 6 of those 7 known escape genes displayed male-biased eQTLs in our analyses. This confirms that some X-linked genetic loci may contribute to shaping sex dimorphisms in both males and females through multiple distinct mechanisms. Interestingly, *GYG2* showed significant eQTL effects with concordant direction across both males and females, but with a larger effect estimate in males (**Fig.2F**), while the other male-only eQTLs showed near zero effect estimates in females (**Table-S4**). This suggests that expression from the second allele in heterozygous females may have potentially attenuated or negated the relative expression effect of the *cis*-regulatory variant compared with hemizygous males. Among the 8 independent PAR eQTLs, one, *VAMP7* in astrocytes, displayed male-only sex heterogeneity at FDR-P_Sex-Het_<0.05 (**Table-S5**), for a total of 10 sex-heterogeneous eGenes at FDR-P_Sex-Het_<0.1 across PAR and non-PAR regions. The relative paucity of sex-biased eGenes at FDR-P_Sex-Het_<0.1 (10/212, 4.7%) is consistent with similar prior QTL studies and likely reflects power challenges to resolve sex-biased QTLs^30–33^, such that future, larger studies will be needed to expand insights into sex-biased X-linked QTLs. Despite this limitation, it should be noted that 30 QTLs corresponding to 28 eGenes reached significance criteria only in our sex-aware analyses (male, female, eXCI), emphasizing the importance of expanding X chromosome analyses beyond standard non-sex-stratified rXCI models.

### Sex-biased X-linked expression is more female-upregulated and shows cell-type heterogeneity

Although assessing evidence for sex-biases or eXCI from QTLs is an important strategy to elucidate sex-dependent gene regulatory effects, various factors, biases, and power limitations may decouple genetics from sex-specific molecular changes. To gain additional insight, we directly assessed differential expression of genes (DEG) across females and males within each of the 7 major brain cell-types (**Fig.3A-G**; **Table-S7**). Between 5 and 50 sex-differentially-expressed genes (sex-DEGs) were identified per cell-type at FDR-P<0.05, corresponding to 7-12% of X-linked genes tested (**Fig.3H**). These signals were robust in sensitivity analyses that adjusted for cognitive diagnosis suggesting the sex-DEGs were not driven by including participants with dementia (**Fig.S3**, Spearman’s ρ=0.998). In total, significant sex-DEGs were observed for 226 cell-type-gene pairs, corresponding to 101 unique genes, of which the majority were female-upregulated (N=164, 72.6%; 67 unique genes). Out of these 101 genes, 44 were significant sex-DEGs across more than one cell-type, including core regulators of X-chromosome inactivation such as *XIST*, *TSIX*, *JPX* and *FTX*^34^, while 57 reached significance criteria in only one cell-type (**Fig.S4**). In contrast to the eQTL findings, most of the 67 female-upregulated genes (49/67, 73.1%) overlapped previously reported escape genes (**Table-S6**)^14^, supporting the sex-DEG approach appeared well-powered to detect potential novel escape genes (cf. later results). While our larger number of female-upregulated genes aligns with expectations for X-linked escape mechanisms, we interestingly still observed 62/226 (27.4%) sex-DEGs (35 unique genes) to be male-biased, supporting other factors such as sex hormone regulation contribute to shaping sex-biased gene expression on the X chromosome. Notably, across all sex-DEGs, only *NLGN4X* was observed in multiple cell-types with discordant effects, showing female-upregulation in neurons and oligodendrocyte progenitor cells (OPCs) versus male-upregulation in oligodendrocytes. Rare genetic variation near *NLGN4X* was recently linked to AD where the genetic signal displayed evidence of eXCI^18^, suggesting that a potential mechanism for *NLGN4X* in AD may be specific to neurons or OPCs.

**Figure 3.**
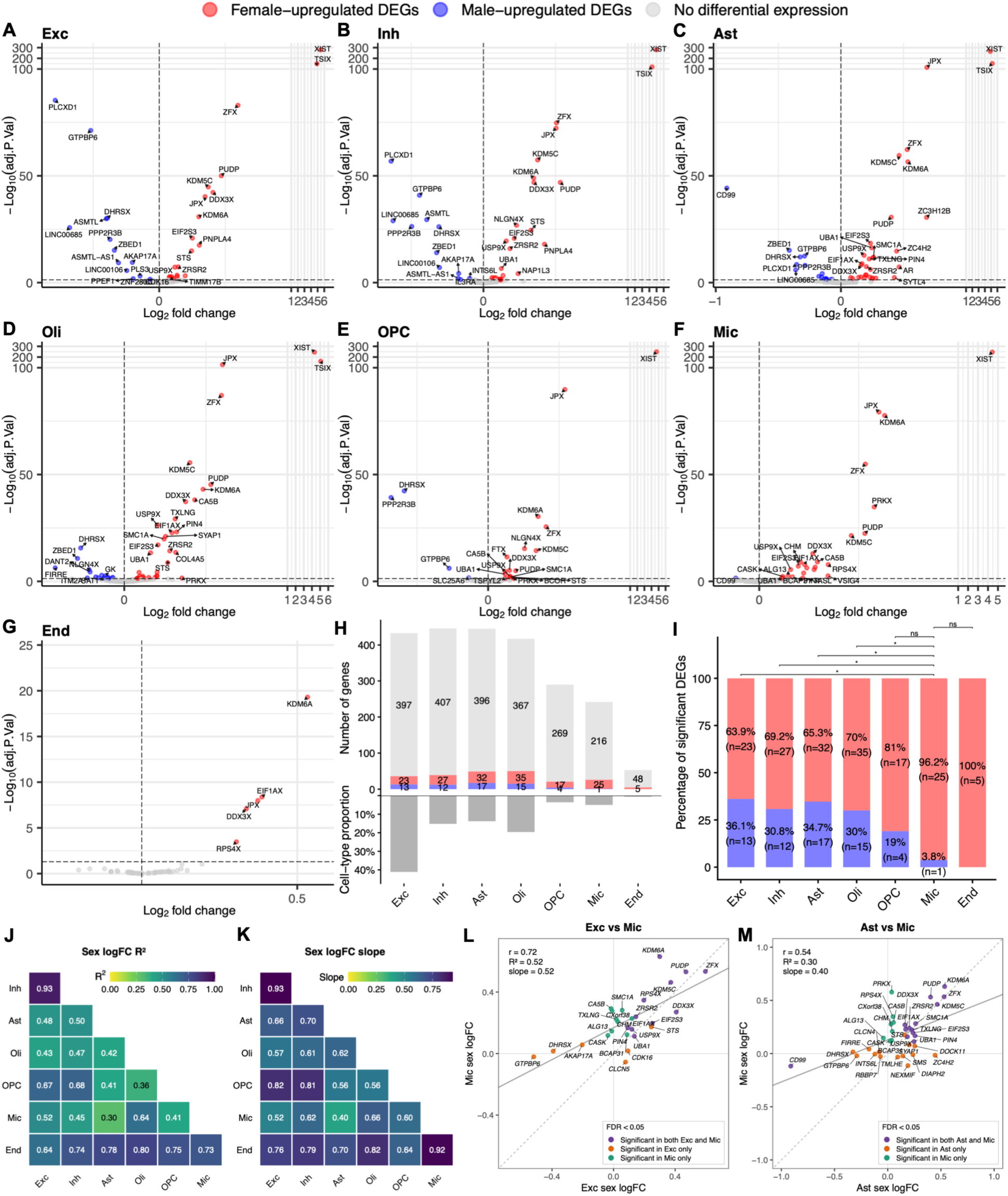
Shared and distinct sex differential expression of X-linked genes (sex-DEG) across cell-types. **A-G)** Volcano plots show cell-type-specific sex-DEG findings across seven brain cell types, showing female-upregulated (red) and male-upregulated (blue) genes passing FDR<0.05. **H)** Top. Total number of genes tested in sex-DEG analysis per cell-type (female-upregulated, red; male-upregulated, blue). Bottom. Cell-type proportions. Note no direct correspondence between number of significant sex-DEGs and cell-type proportions. **I)** Proportion of female-upregulated DEGs (red) versus male-upregulated DEGs (blue) within each cell-type. Note overall larger number of female-biased genes, consistent with expectation for eXCI. Chi-square tests further showed microglia had a significantly higher proportion of female-upregulated DEGs than excitatory neurons, inhibitory neurons, astrocytes, and oligodendrocytes (FDR<0.05). **J-K)** Comparison of effect sizes of significant sex-DEGs between cell-type pairs, showing **J)** R2 and **K)** slope. Note microglia are highly divergent from other cell-types. **L-M)** Scatter plots showing significant sex-DEG log_2_FC values across example cell-type pairs: **L)** Excitatory neurons versus microglia and **M)** Astrocytes versus microglia. Note for (J-M), core regulators of X chromosome escape regulation (e.g. *XIST*) were excluded (cf. Methods & Figure-S6 for sensitivity analyses). *Abbreviations: Exc, excitatory neurons; Inh, inhibitory neurons; Ast, astrocytes; Oli, oligodendrocytes; OPC, oligodendrocyte progenitor cell; Mic, microglia; End, endothelial cells; logFC*, log_2_-fold change.

Further, and again in contrast to the eQTL findings, the proportion of significant sex-DEGs did not follow the same pattern of cell-type abundance, showing relatively higher numbers of sex-DEGs in glial compared to neuronal cell-types (**Fig.3H**). Additionally, chi-square tests showed microglia were particularly enriched for female-upregulated genes relative to other cell-types, having only 1 male-upregulated gene (**Fig.3I**). Comparing the correlation of log_2_-fold changes (log_2_FC) for significant sex-DEGs across cell-types showed high consistency between excitatory and inhibitory neurons, revealing an overall divergence for microglia, astrocytes, and oligodendrocytes (findings from endothelial cells should be considered cautiously given the relative paucity of available genes) (**Fig.3J-K**; **Fig.S5**). This is relevant considering the increasing experimental and genetic evidence that microglia, astrocytes, and oligodendrocytes appear to play important roles in sex differences in AD and that microglial inflammation may disproportionately impact female AD pathogenesis^35–38^. Notably, in the comparison of microglia with excitatory neurons (**Fig.3L**), *KDM6A* showed a 2.1 times larger female-upregulated effect in microglia. Higher *KDM6A* levels in the brain have been linked to resilience to AD^39^, while microglial deletion of *KDM6A* in female mice was shown to confer protection against multiple sclerosis^40^, a disease that like AD has higher female prevalence. This suggests that future studies on the relation of *KDM6A* with AD should specifically interrogate *KDM6A*’s role in microglia. Further, when comparing astrocytes to microglia (**Fig.3M**), *CD99*, a gene that falls in PAR1, which is shared across the X and Y chromosomes, notably showed a 7.7 times larger male-upregulated effect in astrocytes (it was also the most significant male-upregulated gene in astrocytes; **Fig.3C**). A recent cell-type specific study using data overlapping with ours similarly found that astrocyte CD99 signaling was unique to males and linked to AD^25^, while another prior study, through cell-chat analyses, showed interaction of astrocytic *CD99* with microglial *PILRA* (a core AD risk gene) can inhibit inflammatory responses in AD^41^. *CD99* was also reported among genes showing greater silencing near PAR boundaries, which could explain its male-biased upregulation^28^. Overall, the sex-DEG versus sex-informed eQTL approach appeared better powered to identify sex-biased X-linked genes, revealing cell-type heterogeneity and candidate genes that may be relevant to sex-biases in AD and other neurodegenerative disorders.

### X-linked female-biased and escape genes are enriched for Alzheimer’s disease relevance

We next investigated if our female-only/female-biased eQTL (FDR-P_Sex-Het_<0.1) and female-upregulated sex-DEG genes (FDR<0.05; **Fig.2-3**) were specifically relevant to female biases in neurodegeneration. For each cell-type, we classified genes into three groups: our female-biased genes, known eXCI genes not overlapping this set, and all remaining genes (**Fig.4A**; cf. **Methods**). Given the broadly reported female biases in AD^9^, we focused on assessing gene set importance for AD, using enrichment testing with external AD relevance scores from GeneCards (**Fig.4B, Table-S8**). In every cell-type, both our female-biased genes and other known eXCI genes respectively displayed significantly higher scores compared to the remaining tested genes (except our female-biased gene set in microglia which trended towards significance, FDR-P=0.08). Notably, compared to the remaining genes, our female-biased gene sets always displayed the highest differences in AD relevance scores (Cliff’s δ=0.23-0.39) relative to other known eXCI genes (Cliff’s δ=0.21-0.27). For these analyses, we excluded canonical X-inactivation regulators (cf. **Methods**), but results were consistent when including them (**Fig.S7A**). We did not compare our female-biased gene sets with other known eXCI genes given reduced power to do so, such that the larger AD importance of our female-biased genes should be considered as a suggestive finding. Similarly, apparent differences in enrichment of AD relevance between cell-types should be interpreted cautiously, as per-cell-type female-biased gene sets were relatively small and shared many genes. In contrast to AD, PD displays male-biased disease prevalence and enrichment analyses with PD relevance scores from GeneCards showed much less striking relevance of our sex-biased or known escape genes (**Fig.S7B, S8**). This may relate to our snRNAseq data including AD but not PD patients, yet our sensitivity analyses showed QTL and DEG findings were not affected by diagnosis (**Fig.S2-3**).

**Figure 4.**
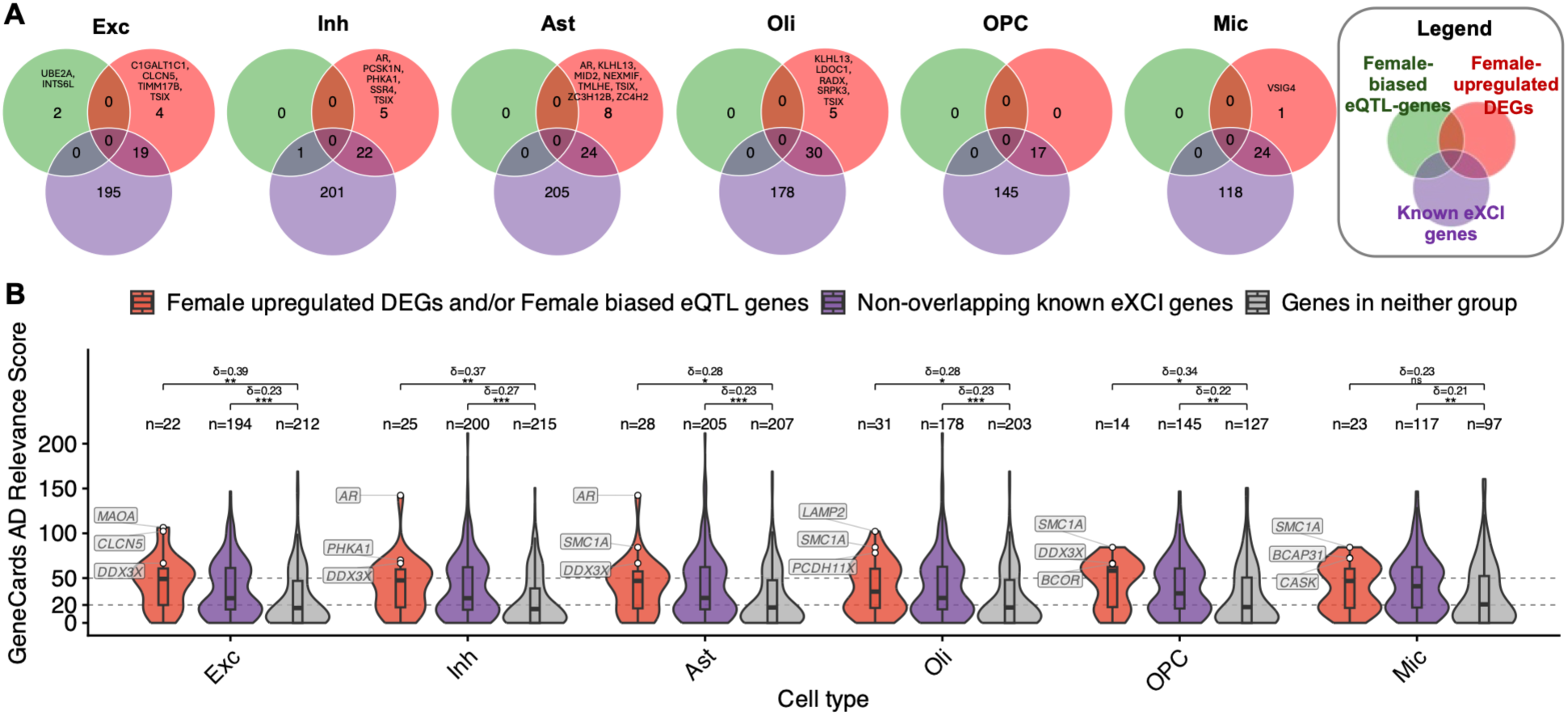
Overview of X-linked genes with female-biased eQTLs or expression levels, known escape status, and their Alzheimer’s disease relevance. **A)** For each cell-type, Venn diagrams show the overlap among female-only or female-biased eQTL genes (FDR-P_Sex-Het_<0.1; green), female-upregulated sex-DEGs (FDR<0.05; red), and previously reported genes that escape X-chromosome inactivation (eXCI genes; purple). Only genes tested in the eQTL analysis for the corresponding cell-type were included. Genes identified in addition to the previously reported eXCI genes are labeled. B) Violin plots of GeneCards AD relevance scores across three gene sets: female-upregulated sex-DEGs and female-biased eQTL genes; non-overlapping previously reported eXCI genes; and genes in neither group, restricted to genes tested in the given cell type. Cliff’s delta indicates the effect size of the between-group difference. Within each cell type, the top three female-upregulated DEGs or female-biased eQTL genes are labeled. Asterisks indicate significance levels (*, FDR<0.05; **, FDR<0.01; ***, FDR<0.001; ns, not significant). Note for (B), core regulators of X chromosome escape regulation (e.g. *XIST*) were excluded (cf. Methods & Figure-S7 for sensitivity analyses).

Beyond general enrichment patterns, we notably observed 20 female-biased genes not previously reported as escape genes (**Fig.4A**). Among these, the androgen receptor gene (*AR*), was top ranked for AD relevance by GeneCards and notably reached female-biased significance in astrocytes and inhibitory neurons (**Fig.4A**; **Table-S7**). The effect was by far strongest in astrocytes (log_2_FC=0.46, FDR=4.8e-8) compared to inhibitory neurons (log_2_FC=0.12, FDR=0.04), while showing small, non-significant female-upregulated effects in excitatory neurons (log_2_FC=0.10, P=0.13) and OPCs (log_2_FC=0.08; P=0.26) and was not sufficiently expressed for testing in other cell-types. This may point to an underappreciated role of androgens in female biology and AD pathogenesis notably within astrocytes. Another gene with high AD relevance was *MAOA*, which encodes mitochondrial enzyme monoamine oxidase A. Prior work showed elevated *MAOA* levels in the frontal cortex of AD patients, as well as a correlation with amyloid-β plaque and tau burden in the anterior cingulate cortex of AD patients^42,43^.

### Cell-type-specific causal inference prioritizes X-linked genes for neurodegenerative diseases

Finally, we integrated our novel X chromosome QTL resource with recent X-chromosome-wide association studies (XWAS) of four neurodegenerative disorders – AD^18^, PD^19^, MSA^21^, and LBD^20^ – to prioritize candidate causal genes within respective cell-types through *in silico* functional genomics analyses. Specifically, we first performed cell-type-specific transcriptome-wide association studies (cTWAS) to identify genes whose genetically predicted expression in respective brain cell-types was associated with disease risk. For those cTWAS gene discoveries passing FDR<0.05 (**Fig.5A**; **Table-S9,** cf. **Methods**), we additionally performed genetic colocalization (COLOC) and summary-based mendelian randomization (SMR) as the primary strategy to prioritize the most robust candidate genes (cf. **Methods**; **Table-S10**). Eleven cTWAS gene discoveries across 4 unique loci and 3 diseases were prioritized for having COLOC (PP4>0.6) or SMR support in at least one cell-type (**Fig.5B**; none were observed in LBD). While cTWAS alone often nominated multiple cell-types for a given gene, after including COLOC and SMR evidence, only 5 genes remained prioritized in multiple cell-types, while 6 prioritized genes appeared cell-type specific (**Fig.5B**). For these 11 prioritized genes in their respective cell-types, we finally compared evidence of sex-biased expression (i.e. reaching at least nominal significance on sex-DEG) to the XWAS model or sex stratum that drove the cTWAS signal in the respective diseases (**Fig.5B**; evidence of sex-biased eQTL effects were not considered given limited power, cf. earlier sections). For the 5 genes prioritized across multiple cell-types, we summed together concordant support from SMR, COLOC, and sex-DEG to nominate the most likely causal cell-type where possible. Across these layers of information, we synthesized insights to nominate top genes, cell-types, and sex-informed mechanisms beyond insights from the prior XWAS efforts. Locus zoom plots for all 11 genes across multiple cell-types and disease associations are provided in **Fig.S9-19**.

**Figure 5.**
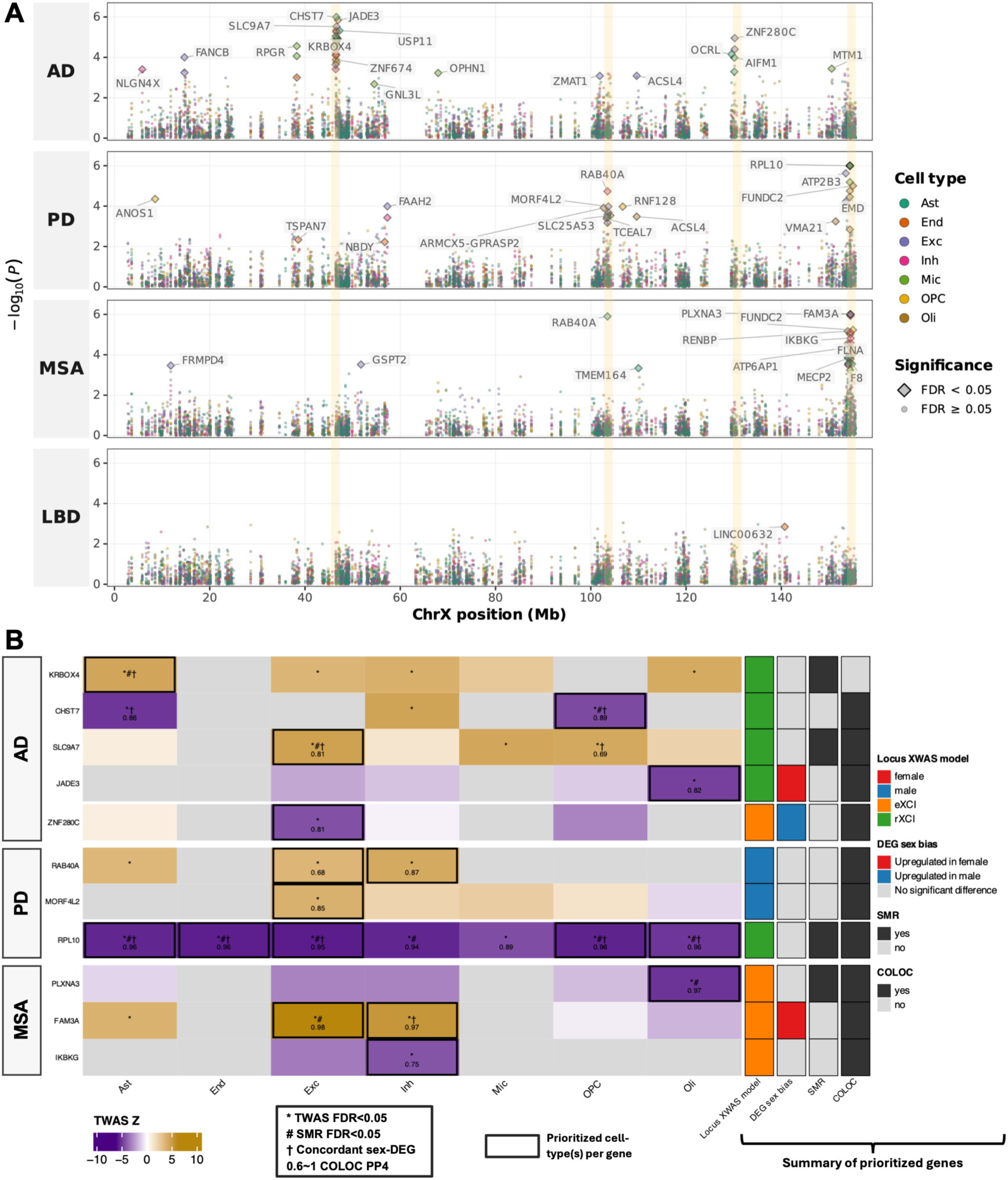
X-chromosome-wide cell-type-specific candidate gene prioritization for neurodegenerative diseases. **A)** Cell-type-specific transcriptome-wide association study (TWAS) results across four neurodegenerative diseases: Alzheimer’s disease (AD), Parkinson’s disease (PD), multiple system atrophy (MSA), and Lewy body dementia (LBD). Each point is colored by cell type; significant TWAS signals (FDR<0.05) are shown as diamonds and labeled with the corresponding gene. Yellow shaded bands indicate **(B)** the prioritized loci additionally supported by summary-data-based Mendelian randomization (SMR) or colocalization (COLOC). The heatmap is colored by TWAS Z-score, with gold and purple respectively indicating increased and decreased gene expression being associated with disease risk. For genes passing cTWAS FDR<0.05 (asterisk), the corresponding cells additionally display evidence of SMR (#, FDR_SMR<0.05 and HEIDI-P-value>0.05), COLOC support (PP4>0.6), or sex-DEG findings were concordant with the underlying XWAS model (†; cf. results and methods for definitions). Black boxes represent the cell-type with the strongest overall prioritization across these multiple streams of evidence. The right section provides a summary of gene prioritization evidence, showing the representative disease XWAS model at the locus, and any SMR, COLOC, or sex-DEG support.

The first AD locus, which included *KRBOX4*, *CHST7*, *SLC9A7*, and *JADE3*, was driven by an rXCI XWAS signal. *JADE3* was only supported by COLOC and showed nominally significant female-biased expression, which is inconsistent with the XWAS rXCI signal underlying the locus. Across the 3 remaining genes, none showed sex biases in sex-DEG analyses, which aligns with the underlying rXCI model. *SLC9A7* in excitatory neurons and *CHST7* in oligodendrocyte progenitor cells were the only genes and cell-types supported by both COLOC and SMR, making these the most promising candidates at the locus. The *SLC9A7* locus was already reported in our prior AD XWAS, where we nominated *SLC9A7* as the candidate causal gene with strong ties to amyloid pathology (cf. Belloy *et al*. 2024 for mechanistic discussion)^18^. Here, we further resolve that the association of *SLC9A7* with AD is most likely mediated within excitatory neurons and corroborate the effect appears shared across both males and females. At the second AD locus, only *ZNF280C* was prioritized. Again, this locus and gene were previously prioritized^18,44^, but we here further resolved the likely causal cell-type to be excitatory neurons. However, while the underlying XWAS signal indicated an eXCI effect, *ZNF280C* showed male-upregulated expression at FDR-P=0.028 in excitatory neurons, suggesting future studies will be needed to resolve this ambiguity. For PD, one locus, containing *RAB40A* and *MORF4L2*, was not previously identified^19^. Both genes were prioritized within excitatory neurons and *RAB40A* additionally in inhibitory neurons. The underlying XWAS model indicated a male-biased effect at this locus, but this was not evident even at nominal significance in the sex-DEG findings for the nominated gene-and-cell-type pairs, such that future efforts should validate if a true male-biased genetic effect exists. Notably, *MORF4L2* appears relevant to PD from recent knowledge graph analyses of monogenic PD forms^45^. At the *RPL10* locus, where the prior PD XWAS already nominated *RPL10*^19^, all 7 cell-types were nominated for *RPL10* with no clear evidence in favor of one cell-type. Assessment of sex-DEG findings confirmed alignment of the underlying rXCI XWAS signal with no observable sex-biased effects for *RPL10*, except in microglia and inhibitory neurons where *RPL10* showed evidence of female-biased expression, suggesting these are less likely to be the causal cell-types. Notably, *RPL10* lies within the same broad Xq28 locus implicated by the MSA XWAS and cTWAS, yet the two diseases pinpoint distinct genes, cell-types, and X-linked dosage models. The prior MSA XWAS identified a female-biased signal and used GTEx eQTL colocalization to prioritize *PLXNA3* in cervical spinal cord and *FAM3A* in hypothalamus^21^. Through extended analyses (cf. Methods), we determined this MSA signal appears more significant under the eXCI model. We further implicated *PLXNA3* in oligodendrocytes, *FAM3A* in both inhibitory and excitatory neurons, and *IKBKG* in inhibitory neurons. The nomination of *PLXNA3* in oligodendrocytes is of notable interest given that α-synuclein expression specifically in oligodendrocytes appears relevant to MSA^46^. While we did not observe evidence of XCI escape for *PLXNA3*, recent work showed that *PLXNA3* may show aging-induced escape, which would align with the underlying eXCI model at this locus^47^. Further, while *FAM3A* had both COLOC and SMR support in excitatory neurons versus only COLOC support in inhibitory neurons, sex-DEG findings indicated a nominal female-biased effect for *FAM3A* only in inhibitory neurons which aligned with the underlying eXCI model. *FAM3A* plays a role in mitochondrial function which in turn may be relevant to MSA^48^. In conclusion, *FAM3A and PLXNA3* are the most promising candidate genes at this MSA locus, with our findings providing novel insights into their likely causal cell-types and sex-biased mechanisms.

## DISCUSSION

We present a first-in-kind cell-type-specific map of X-chromosome gene regulation in the aged human brain, leveraging a large sample of 423 individuals. Our sex-aware analyses showed stronger significance for 18.9% of non-PAR X-linked eQTLs while also expanding eQTL discovery by 6% relative to non-sex-aware analyses, revealed novel sex-biased eQTLs, and found important cellular heterogeneity in sex-biased expression of X-linked genes with notable enrichment of female-upregulated genes in microglia, which is consistent with growing evidence that immune-related cell-types and mechanisms are important to sex differences in neurodegeneration and brain aging^9,36,49–51^. We additionally showed that female-upregulated and escape genes are enriched in relevance for Alzheimer’s disease, but less so for Parkinson’s disease, consistent with the sex-biased prevalences of these diseases. Finally, by integrating our resource with XWAS of four neurodegenerative diseases, we prioritized 11 candidate causal genes annotated with their putative sex-biased or sex-shared effects.

Primary limitations of this study include power challenges in detecting sex-biased eQTLs, a focus on major brain cell-types rather than subtypes (this choice was made because of anticipated power limitations), the European ancestry of the ROSMAP cohort which limits generalizability across populations, and the use of an AD-enriched dataset from a single brain region (DLPFC) as a proxy for the aged brain. While models adjusting for diagnostic groups showed high consistency of QTL and DEG findings, other brain regions and cohorts enriched for other diseases will obviously be important to expand insights of X-linked gene regulation in neurodegenerative disease broadly. Focus points for future studies should thus be expanding sample sizes in more diverse populations and diseases. Extended studies may also explore allele-specific expression analyses for detection of escape genes and seek to evaluate effects of X inactivation skewing, age-related X reactivation, or other mechanisms relevant to sex chromosomes and their interaction with sex hormones^11,52^. It should finally be noted that our causal gene inference results pertain to *in silico* analyses, thus requiring experimental validation, and that we focused on the current cell-type specific eQTL data to nominate genes, but other genes or mechanisms could additionally be prioritized through different tissue contexts and molecular QTL types.

In summary, our findings add to the growing consensus that X-linked gene regulation plays an important role in shaping sex differences in the aged brain and neurodegenerative disease risk. Our resource will importantly support future studies of the X chromosome and contributes to accelerating sex-informed precision medicine in neurodegeneration.

## METHODS

### Study cohort

Brain samples with accompanying single-nucleus RNA-sequencing (snRNA-seq) and whole genome sequencing (WGS) data were obtained from the Religious Orders Study (ROS) and the Memory and Aging Project (MAP), two longitudinal cohorts of aging analyzed jointly as ROSMAP^53^. Both studies were approved by the Institutional Review Board of Rush University Medical Center, and all participants provided written informed consent, an Anatomical Gift Act, and a repository consent. Cohort design and recruitment are described in detail before^24,53^. 424 ROSMAP participants were retained after intersecting snRNA-seq and WGS data, with 1 subject being excluded during quality control for a total of 423 participants (288 female and 135 male) for X-chromosome centric analyses. Cell-type-specific demographics of included individuals after quality control are summarized in **Table S1**.

### Single-nucleus RNAseq processing

The snRNA-seq data were generated and processed by Fujita et al^24^. Nuclei from frozen DLPFC tissue were profiled on the 10x Genomics Chromium platform in pools of eight participants, and each nucleus was assigned to its donor by genotype-based demultiplexing against the ROSMAP WGS data. After removal of low-quality nuclei and doublets, nuclei were classified into major cell-types, of which seven had sufficient data for eQTL mapping (excitatory neurons, inhibitory neurons, astrocytes, oligodendrocytes, oligodendrocyte progenitor cells, microglia, and endothelial cells). Pseudobulk expression was then derived per cell-type by aggregating UMI counts per gene across all nuclei of that cell-type within each individual, excluding individuals with fewer than ten nuclei of a given cell-type.^24^

Following Fujita et al.^24^, pseudobulk counts were normalized (for all samples across both sexes) using the trimmed mean of M-values method in edgeR^54^, log2 counts per million were computed with the voom function of limma^55^, and genes with a mean log2 counts per million below 2 were removed. Batch effects were corrected with the ComBat function of sva while preserving the sex effect, and expression values were quantile normalized. In addition, gene symbols were standardized with the HGNChelper package to update alias symbols and remove duplicated symbols^56^, and analyses were restricted to X-linked genes defined from the GENCODE v48 basic annotation.

### X-chromosome genotype quality control

All genetic quality-control steps were performed with PLINK (v1.9 and v2.0)^57^. WGS data from ROSMAP donors were intersected with the 424 subjects for which pseudobulk snRNAseq was available^24^. Autosomal data was processed to identify relatedness and ancestry, confirming all subjects were of European ancestry and unrelated. X chromosome genetic data was extracted and an initial genotyping-rate filter (genotyping rate >95%, minor allele count ≥1) was applied across all samples^53^. Variants were then required to have genotyping rate >95% within cognitively normal and cognitively impaired individuals respectively, and within each sex respectively. Further, variants showing differential missingness by diagnosis (P<1e^-5^) or sex (P<1e^-10^) were removed. An additional Fisher exact test was used in cognitively normal individuals to remove variants with genotype distributions deviating across sexes (P<1e^-5^). Hardy-Weinberg equilibrium was evaluated in cognitively normal individuals of both sexes using the X-chromosome exact test in PLINK2^58^, and variants with P<1e^-5^ were removed. In the non-PAR region, male heterozygous hard calls were set to missing, after which samples exceeding a genotype missingness rate of >2% were removed. Three additional filters were applied using gnomAD v3.1.1 to remove variants located in gnomAD low-complexity regions, variants flagged as non-PASS in gnomAD, and variants whose control-sample minor allele frequency differed from the gnomAD non-Finnish European frequency by more than 10%^59^. Finally, minor allele frequencies were computed in the combined-sex, male-only, and female-only groups, and a variant was retained if it had a MAF ≥0.01 in all three groups. In total, 238,428 non-PAR and 5,358 PAR variants (243,786 in total) were retained. Of these, 224,853 fell within the *cis*-window (± 1Mb of transcription start site) of at least one tested gene and were used for *cis*-eQTL mapping.

### Cell-type-specific *cis*-eQTL mapping

*Cis*-eQTL mapping was performed with PLINK 2.0^57^, testing X-chromosomal variants against the expression of each testable gene in the corresponding cell-type, with covariates standardized to unit variance. For each cell-type, covariates comprised age at death, postmortem interval, study (ROS versus MAP), the total number of detected genes, three genotype principal components (PCs), and 30 expression PCs^24^. Genotype PCs were computed from autosomal variants after LD pruning, and expression PCs were derived from the 1,000 most variable non-X-linked genes. Missing postmortem-interval values were imputed with the cell-type median. In the combined-sex analyses, sex was additionally included as a covariate. Non-PAR and PAR variants were analyzed separately. For non-PAR variants, four analyses were conducted: a random-X-inactivation model (rXCI) and an escape model (eXCI), each estimated in the combined-sex sample, as well as female-only and male-only analyses. The rXCI and eXCI settings were implemented through PLINK’s X-chromosome dosage coding for males. Under rXCI (--xchr-model 2), which assumes full dosage compensation, hemizygous males were coded as 0/2. Under eXCI (--xchr-model 1), which assumes no dosage compensation, males were coded as 0/1. The male-only analyses also used 0/1 coding. For PAR variants, which are diploid in both sexes, three analyses were conducted under standard additive coding: combined-sex, female-only, and male-only. For each gene, *cis* variants were defined as those within 1 Mb of the transcription start site, with the transcription start site in GENCODE v48^60^. As a sensitivity analysis, cis-eQTL mapping was repeated with cognitive diagnosis at death (no cognitive impairment versus impaired) added as a covariate, and effect estimates from the primary and adjusted analyses were correlated across all tested variant-gene-cell-type-model combinations and across those significant in both analyses. Although our study focused on *cis*-regulation, cell-type-specific *trans*-eQTL summary statistics for all X chromosome variants with each respective X-linked gene were generated and provided as a resource (cf. data availability).

### Identification of independent eQTL signals

Significant *cis*-eQTLs were identified within each cell-type and analysis model or stratum, using a two-step false discovery rate (FDR) procedure^24^. In the first step, the *cis* variant P-values for each gene were adjusted for the number of *cis* variants tested at that gene using Bonferroni correction, and the smallest adjusted value was taken as the gene-level P-value. In the second step, gene-level P-values were corrected across all tested genes within the respective cell-type using the Benjamini-Hochberg procedure, and the gene-level threshold corresponding to FDR<0.05 was determined. A *cis* variant was declared significant if its gene-level-adjusted P-value did not exceed this threshold. Independent signals were resolved by linkage disequilibrium (LD) clumping using ROSMAP X-chromosome WGS data as the reference panel (clumping window 1Mb, r² threshold 0.1). Clumping was first performed for each respective gene, cell-type, and model or sex. Because the same or neighboring variants could reach significance under more than one model, the significant lead variants across all models were then combined, within each gene and cell-type, in a second round of LD clumping, ranking variants by their smallest P-value across models. The variants surviving this second round constituted the final set of independent *cis*-eQTL signals.

### Sex-heterogeneity testing of X-linked *cis*-eQTLs

To assess whether X-linked *cis*-eQTL effects differed between sexes, sex-heterogeneity testing was performed for the independent *cis*-eQTL signals in the non-PAR (**Table-S4**) and PAR (**Table-S5**) regions. For each independent signal, sex-stratified effect sizes were taken from the female-only and male-only analyses. In the non-PAR regions, because male-only analysis used 0/1 genotype coding, male effect sizes and standard errors were divided by two to place them on the same scale as females (essentially rendering them on the rXCI scale) before sex-heterogeneity testing. Sex heterogeneity was tested with a two-sample z-statistic, Z_Sex-Het_ = (β_male_ − β_female_) / √(SE_male_^2^ + SE_female_^2^), with a two-sided P value (P_Sex-Het_) obtained from the standard normal distribution. P_Sex-Het_ values were corrected with the Benjamini-Hochberg procedure jointly across all tested non-PAR and PAR signals (FDR-P_Sex-Het_). Signals with P_Sex-Het_<0.05 were considered to show potential sex heterogeneity and were further classified based on their sex-stratified results. Signals passing the two-step FDR threshold in only one sex-stratified analysis were labeled “female-only” or “male-only”; all remaining signals were attributed to the sex with the larger absolute effect size and labeled “female-biased” or “male-biased”. This yielded four categories: female-only and male-only (significant in females or males only, respectively) and female-biased and male-biased (stronger in females or males, respectively). Signals with P_Sex-Het_≥0.05 were classified as showing no evidence of sex heterogeneity. A more lenient threshold at FDR-P_Sex-Het_<0.1 was used in addition to the common FDR-P_Sex-Het_<0.05 threshold to summarize findings, consistent with prior studies of sex-biased QTLs^27^.

### Sex differential expression of genes (sex-DEG) analyses

For each cell-type, raw pseudobulk counts were analyzed using edgeR and limma. Lowly expressed genes were first filtered using filterByExpr, followed by trimmed mean of M-values normalization. Normalized counts were transformed with voom and genes with mean log2 counts per million ≤2 were removed^24^. The remaining genes were then reprocessed with voom and fitted using linear models implemented in limma. The primary model tested the effect of sex on gene expression while adjusting for study, postmortem interval, age at death, and batch. Gene symbols were updated using the HGNChelper package ^56^. Within each cell type, genes tested in sex-DEG analyses were the same as those tested in eQTL analyses. Multiple-testing correction was then calculated using the Benjamini-Hochberg false discovery rate procedure across the tested X-linked genes within each cell-type (FDR<0.05). Because the sex-effect coefficient represented female versus male expression, sex-DEGs with positive log_2_FC were classified as female-upregulated, whereas those with negative log_2_FC were classified as male-upregulated. As a sensitivity analysis, the sex differential expression analysis was repeated with additional adjustment for cognitive impairment status, defined from ROSMAP cognitive diagnosis as no cognitive impairment versus any cognitive impairment. Concordance between the primary and sensitivity analyses was assessed by correlating sex-effect log_2_FC estimates across all tested cell-type-gene pairs. When comparing sex-DEG effect estimates across cell-types, master regulators of X-inactivation (*XIST*, *TSIX*, *JPX*, *FTX*, *RLIM*, and *XACT*) were excluded from analyses.

### Curated list of known eXCI-escape genes

Known eXCI genes were curated from twelve previous studies assessing X-chromosome inactivation escape and sex biases^13,14,28,61–69^. For each gene in each study, an escape score was assigned as 0 for genes reported as not escaping, 0.5 for genes reported as variably escaping, and 1 for genes reported as escaping. Scores were summed across the twelve studies, and genes with a total escape score ≥1 were defined as known eXCI genes (**Table-S6**).

### Gene set disease relevance enrichment

AD and PD relevance scores were respectively derived by querying GeneCards with the term “Alzheimer’s disease” or “Parkinson’s disease” and exporting the resulting GeneCards Relevance Scores^70^. Endothelial cells were excluded because few X-linked genes were testable in this cell-type (N=53 versus 242-446 in the others). Within each cell-type, tested genes were partitioned into three groups: (1) female-biased genes identified in this study (female-upregulated DEGs at FDR<0.05 and/or female-only or female-biased eQTL genes at FDR<0.1); (2) known eXCI genes with prior evidence of escape that did not overlap group 1; and (3) all remaining genes tested in that cell-type not belonging to the two prior groups. Disease relevance scores were compared between group 1 and group 3, and between group 2 and group 3, within each cell-type. Normality was assessed for each group with the Shapiro-Wilk test; a two-sided t-test was used when both groups were normally distributed and a two-sided Mann-Whitney U test otherwise, requiring at least three genes per group. Effect sizes were quantified with Cliff’s δ (tie-corrected, range -1 to 1), with positive values indicating higher relevance scores in the female-biased or known escape gene group relative to the remaining genes. P-values were corrected for multiple testing using the Benjamini-Hochberg procedure across all cell types and comparisons (global FDR). Master regulators of X-inactivation (*XIST*, *TSIX*, *JPX*, *FTX*, *RLIM*, and *XACT*) were excluded from the primary analyses.

### Cell-type-specific transcriptome-wide association studies (cTWAS)

X-chromosome-wide association summary (XWAS) statistics were obtained for AD^18^, PD^19^, MSA^21^, and LBD^20^ and covered only the non-PAR regions. Gene expression prediction weights were constructed following the FUSION framework^71^. Within each cell-type, weights were built separately under three strata (combined-sex, female-only, and male-only). For each gene, variants within ±1 Mb of the transcription start site were extracted and the phenotype was the normalized expression of that gene. The covariates were identical to those used in the main *cis*-eQTL analysis, with sex omitted in the female-only and male-only strata. cis-heritability was estimated with GCTA^72^, and predictive weights were fit using PLINK^57^ and GEMMA as implemented in the FUSION weight-computation pipeline. Weights were retained for every gene for which they could be estimated, without applying a *cis*-heritability significance filter. For each disease, four association models were used: a random-X-inactivation model (rXCI) and an escape model (eXCI), both derived from the combined-sex sample, together with female-only and male-only analyses; the eXCI summary statistics were generated by conducting random effects meta-analyses of male XWAS recoded on the 0/1 scale with female XWAS, which effectively approximates eXCI^44,73,74^. Summary statistics were on GRCh38 (lifted over from GRCh37 where required), effect estimates were converted to Z statistics (Z = β/SE), and variant identifiers in the summary statistics were relabeled to match those in the ROSMAP reference (weight files) by chromosome, position, and alleles. Gene-level associations were computed with FUSION^71^, using cohort-specific WGS data as the linkage-disequilibrium reference panel. Each disease association model was paired with weights as follows: the rXCI XWAS was tested against combined-sex weights only, whereas the eXCI, female-only, and male-only XWAS were each tested against two weight sets; the eXCI and female-only XWAS were each tested against combined-sex and female-only weights, and the male-only XWAS against combined-sex and male-only weights. These combinations reflect the sex and model matches in addition to also leveraging the increased power from non-sex-cTWAS weights. Gene-level associations were computed separately for each combination of disease, XWAS model, weight set, and cell type. Within each such combination, associations with FDR<0.05 were considered supported by cTWAS.

### Integration of cTWAS with SMR, COLOC, and sex-DEG insights for gene prioritization

For candidate genes identified by cTWAS, we used support from summary-based mendelian randomization (SMR) or genetic colocalization (COLOC) analyses to prioritize respective genes and cell-types^75,76^. SMR tests whether a disease association and a cis-eQTL signal are consistent with a shared causal variant^75^, using the top cis-eQTL variant as the instrument, and the accompanying HEIDI test flags associations driven by linkage disequilibrium rather than a single shared variant. XWAS and cell-type cis-eQTL summary statistics from all settings (rXCI, eXCI, female, and male) were harmonized to the ROSMAP X-chromosome reference, with male eQTL effects rescaled to the 0/2 dosage scale. SMR was run with ROSMAP X-chromosome genotypes as the LD reference. SMR P-values were FDR-corrected across genes within each disease, XWAS model, and cell-type, and SMR support was defined as FDR-P_SMR_<0.05 with P_HEIDI_>0.05. COLOC was performed with coloc.abf using default priors (p_1_=1×10^-4^, p_2_=1×10^-4^, p_12_=1×10^-5^)^77^ on variants within ±1 Mb of each gene’s transcription start site; COLOC support was defined as PP4>0.6. Pairing of XWAS and eQTL data in SMR and COLOC was consistent with cTWAS analyses (cf. above).

Genes with a significant cTWAS association (FDR-P<0.05) that were additionally supported by SMR or COLOC in the same cell-type were prioritized for further interrogation and review. For each locus, the XWAS model (rXCI, eXCI, female, or male) of the prioritized gene with the strongest cTWAS association was taken as the representative model or sex-bias of the locus. For each prioritized gene, we then asked whether this model matched the sex-DEG result in the prioritized cell-types (nominal P<0.05): an eXCI or female XWAS signal would be expected to coincide with female-upregulated expression, a male signal with male-upregulated expression, and an rXCI signal with no sex difference. When a gene was prioritized in several cell-types, the level of evidence across SMR, COLOC, and sex-DEG was used to further prioritize the most likely causal cell-types.

## Data availability

Data used in the cell-type-specific eQTL analyses are available upon application to:

- ROSMAP whole-genome sequencing, AD Knowledge Portal / Synapse (https://www.synapse.org/Synapse:syn10901595)
- ROSMAP single-nucleus RNA-seq and pseudobulk expression^24^, AD Knowledge Portal / Synapse (https://www.synapse.org/Synapse:syn52335807)

Cell-type-specific X-chromosome *cis* and *trans*-eQTL summary statistics generated in this study are available through the GeneXplore browser (https://genexplore.wustl.edu/) and are also available, together with the cTWAS expression weights, in Zenodo repository (https://doi.org/10.5281/zenodo.23989152).

XWAS summary statistics used in the causal inference analyses are publicly available:

- Alzheimer’s disease (https://github.com/Belloy-Lab/AD_XWAS_Pleiotropy)
- Parkinson’s disease (GWAS Catalog; GCST90104085–GCST90104087)
- Lewy body dementia (GWAS Catalog; GCST90502821–GCST90502823)
- Multiple system atrophy (GWAS Catalog; GCST90558035–GCST90558037)

Reference resources used for variant and gene annotation are publicly available:

- gnomAD v3.1.1 (https://gnomad.broadinstitute.org/)
- GENCODE v48 (https://www.gencodegenes.org/human/)
- dbSNP (https://www.ncbi.nlm.nih.gov/snp/)
- GeneCards (https://www.genecards.org/)
- Previously reported escape genes, curated from published studies (**Table-S6**)

## Code availability

All analysis code is available at https://github.com/Belloy-Lab/X_chr_sceQTL. The following publicly available software was used: PLINK v1.9 and v2.0 (https://www.cog-genomics.org/plink/), FUSION (http://gusevlab.org/projects/fusion/), SMR v1.3.1 (https://yanglab.westlake.edu.cn/software/smr/), and the coloc R package v5.2.3 (https://chr1swallace.github.io/coloc/).

### Acknowledgements

This work was supported by grants from the National Institute on Aging (R00AG075238, M.E.B.), Cure Alzheimer’s Fund (M.E.B.), and Alzheimer’s Association (AARG-24-1027303). This work was also supported by access to equipment made possible by the Hope Center for Neurological Disorders, the Neurogenomics and Informatics Center (NGI: https://neurogenomics.wustl.edu/) and the Departments of Neurology and Psychiatry at Washington University School of Medicine. We thank all study participants and their families as well as the many involved institutions and their staff. The results published here are in whole or in part based on data obtained from the AD Knowledge Portal (https://adknowledgeportal.synapse.org/). The ROS/MAP datasets used in this study were funded by NIH grants U01AG061356, RF1AG057473 and U01AG046152 as part of the AMP-AD consortium.

## Author Contributions

Y.Z. performed data acquisition, designed analyses, performed analyses, interpreted results, and wrote the paper. N.C. curated data, performed analyses, and contributed to analytical design. M.F. and V.M. contributed to data acquisition. M.J., C.P., and C.C. contributed to data acquisition and curated data. A.P.W., T.S.W., and C.C. contributed data. A.R.D. contributed to interpretation of results and reviewed the manuscript. M.E.B. conceptualized and designed the study, performed data acquisition, designed analyses, supervised analyses, supervised work, wrote the paper, and obtained funding.

