## Supplementary Figures S1-S19 for "Brain Cell-Type-Specific X-Chromosome Gene Regulation Reveals Sex-Biased Candidate Genes in Neurodegeneration"

### Supplementary Material

***Supplementary Figures*** .......... **2**

Supplementary Figure S1. Landscape of X-chromosome-wide cell-type-specific eQTLs in the DLPFC, restricted to common variants (MAF ≥ 0.05) .......... 3

Supplementary Figure S2. Comparison of effect size estimates before and after adjusting for cognitive status .. 5

### Supplementary Figures

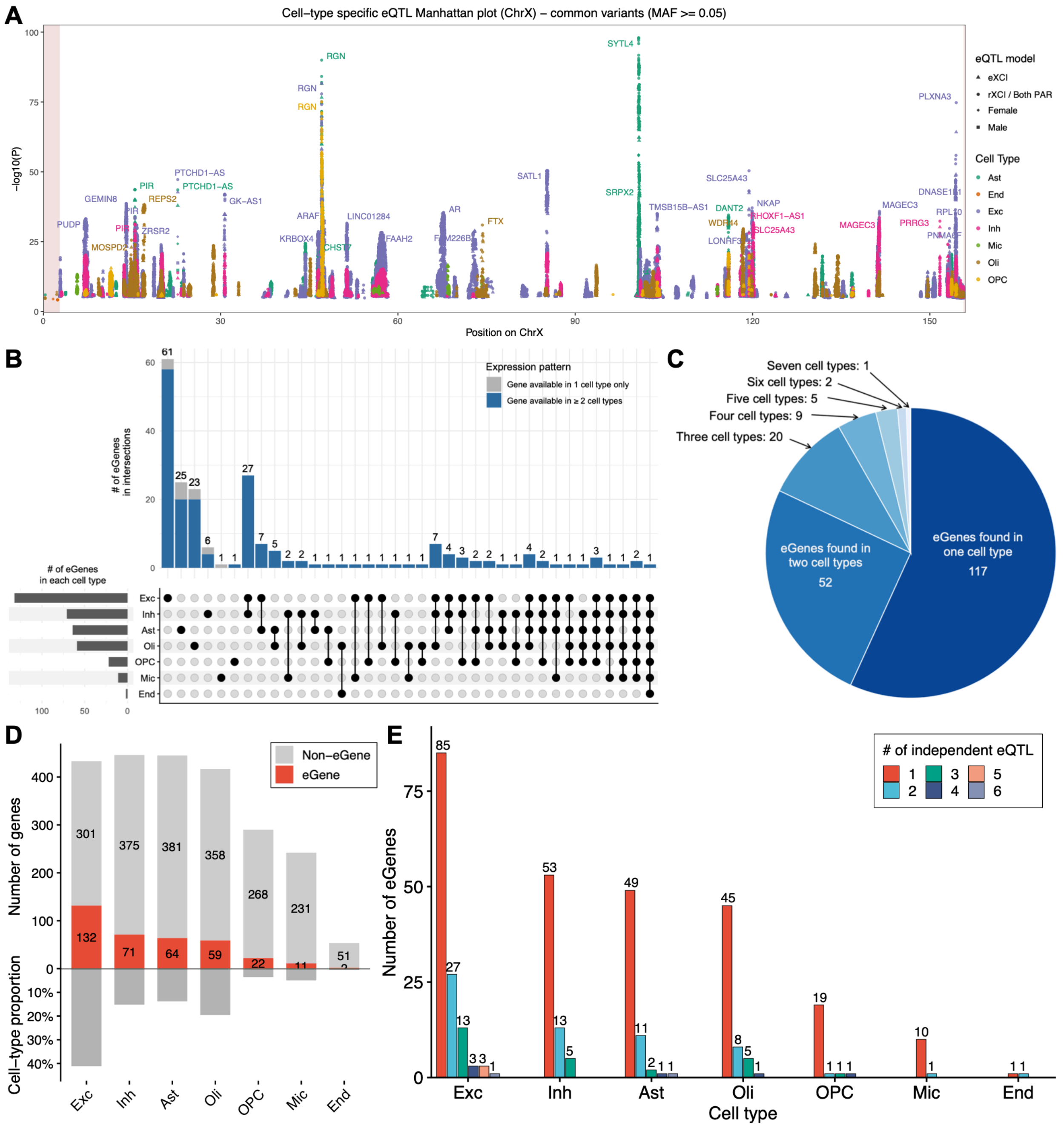

**Supplementary Figure S1. Landscape of X-chromosome-wide cell-type-specific eQTLs in the DLPFC, restricted to common variants (MAF ≥ 0.05).**

This figure reproduces the analyses in Figure 1 after restricting to variants with minor allele frequency (MAF) ≥ 0.05 in all sex groups (sex-combined, female-only, and male-only).

**A** Manhattan plot showing cell-type-specific eQTL associations across the X chromosome. Only associations passing the two-step FDR threshold are displayed. Points are colored by cell type and shaped according to the X-chromosome dosage model used for eQTL testing. The top eGenes (genes with at least one significant eQTL) at each locus are labeled. Shaded regions at the start and end of the X chromosome indicate PAR1 and PAR2, respectively.

**B** UpSet plot showing eGenes unique to or shared across cell types. Vertical bars represent the number of eGenes in each cell-type intersection. Horizontal bars indicate the total number of eGenes per cell type. Bars are colored by expression availability pattern, distinguishing genes available in one cell type only (gray) from those available in two or more cell types (blue).

**C** Distribution of cell-type-specific eGenes (significant eQTL available in one cell type only) and non-cell-type-specific eGenes (significant eQTL in at least two cell types).

**D** Top. Number of genes tested per cell type, with genes classified as eGenes or non-eGenes. Bottom. Cell-type proportions. Excitatory neurons showed a higher number of eGenes consistent with their higher cell-type proportion.

**E** Number of eGenes with different numbers of independent significant eQTL signals in each cell type.

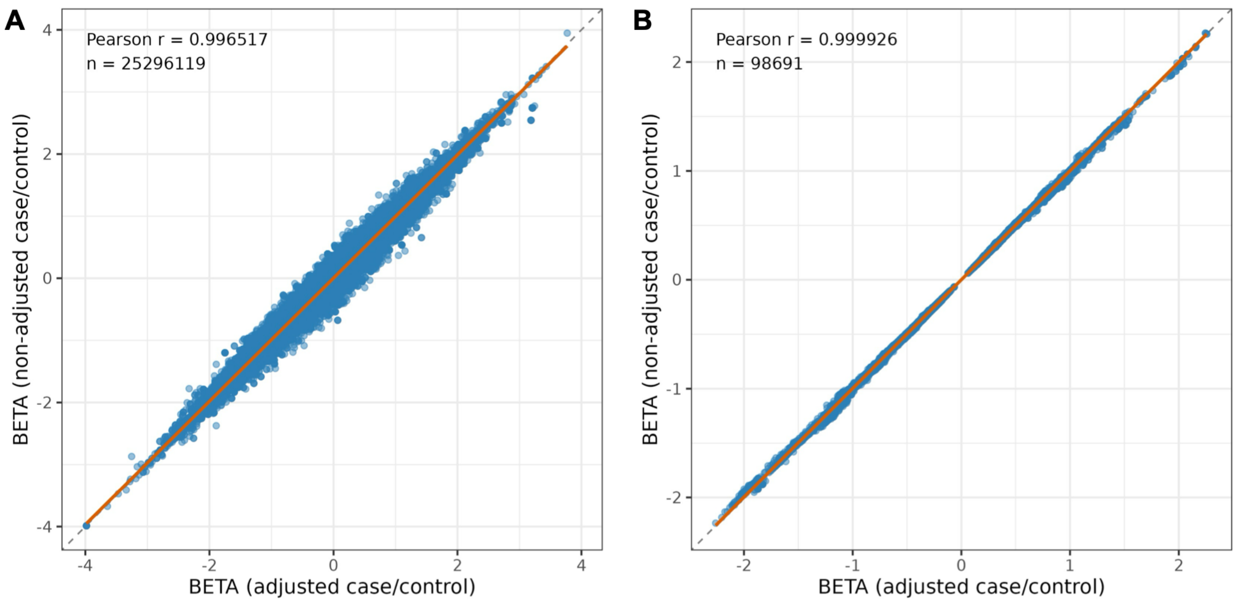

**Supplementary Figure S2. Comparison of effect size estimates before and after adjusting for cognitive status.** Each point is one tested association (variant × gene × cell type × X-chromosome dosage model or sex stratum). Effect sizes (BETA) from models additionally including cognitive status (no cognitive impairment versus any cognitive impairment) as a covariate (x axis) are compared against those from the primary models (y axis). The dashed grey line indicates the identity line ($y = x$) and the orange line the linear regression fit; the Pearson r and number of associations (n) are shown in each panel. Effect sizes were highly concordant between the two models, indicating that adjustment for cognitive status had little influence on the eQTL effect estimates. **A** All tested associations (n = 25,296,119). **B** Associations passing two-step FDR in both analyses (n = 98,691).

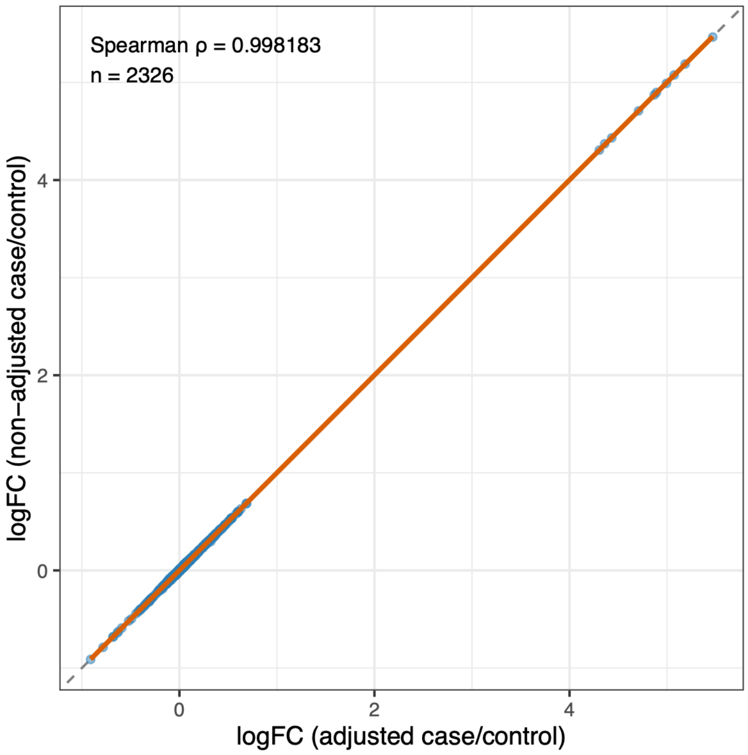

**Supplementary Figure S3. Comparison of differential expression effect sizes (logFC) before and after adjusting for cognitive status.** Each point is one cell-type-gene sex DEG result; the orange line is the identity line (y = x), with the Spearman ρ and number of comparisons (n) shown. Effect sizes were highly consistent between the two models (Spearman ρ = 0.998, n = 2,326), indicating that cognitive status had little influence on the estimates.

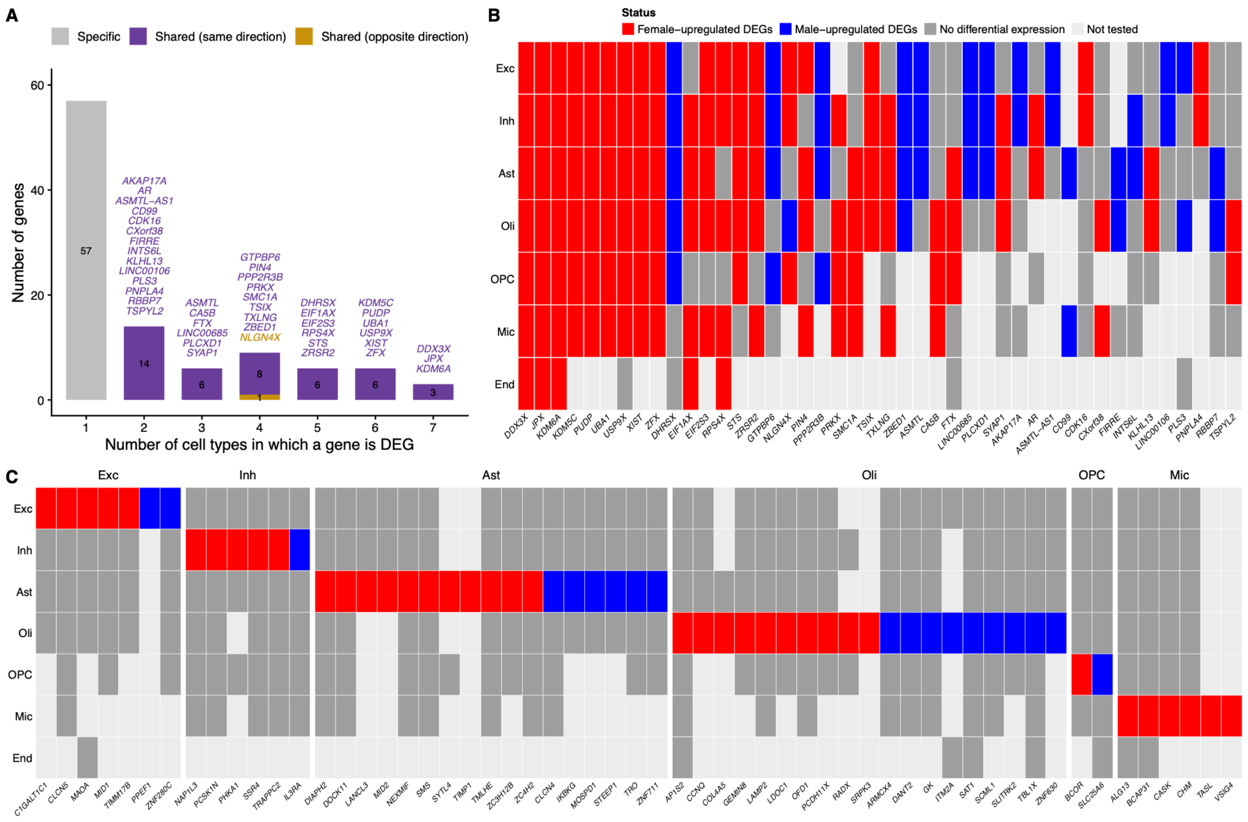

**Supplementary Figure S4. Cell specificity of sex differentially expressed genes (DEGs) on the X chromosome.** **A** Number of sex DEGs detected in a given number of cell types (1–7). Bars are colored by sharing pattern: sex DEGs detected in only one cell type (specific, gray), sex DEGs detected in multiple cell types with the same direction of sex-bias (shared, same direction), and sex DEGs detected in multiple cell types with discordant directions (shared, opposite direction).

**B** Heatmap of shared sex DEGs (DEGs detected in ≥2 cell types). Rows are cell types and columns are genes; each cell is colored by sex-bias status: female-upregulated DEG (red), male-upregulated DEG (blue), no differential expression (gray), or not tested (light gray).

**C** Heatmap of cell-type-specific sex DEGs, grouped by cell type, with the same color scheme as in **B**.

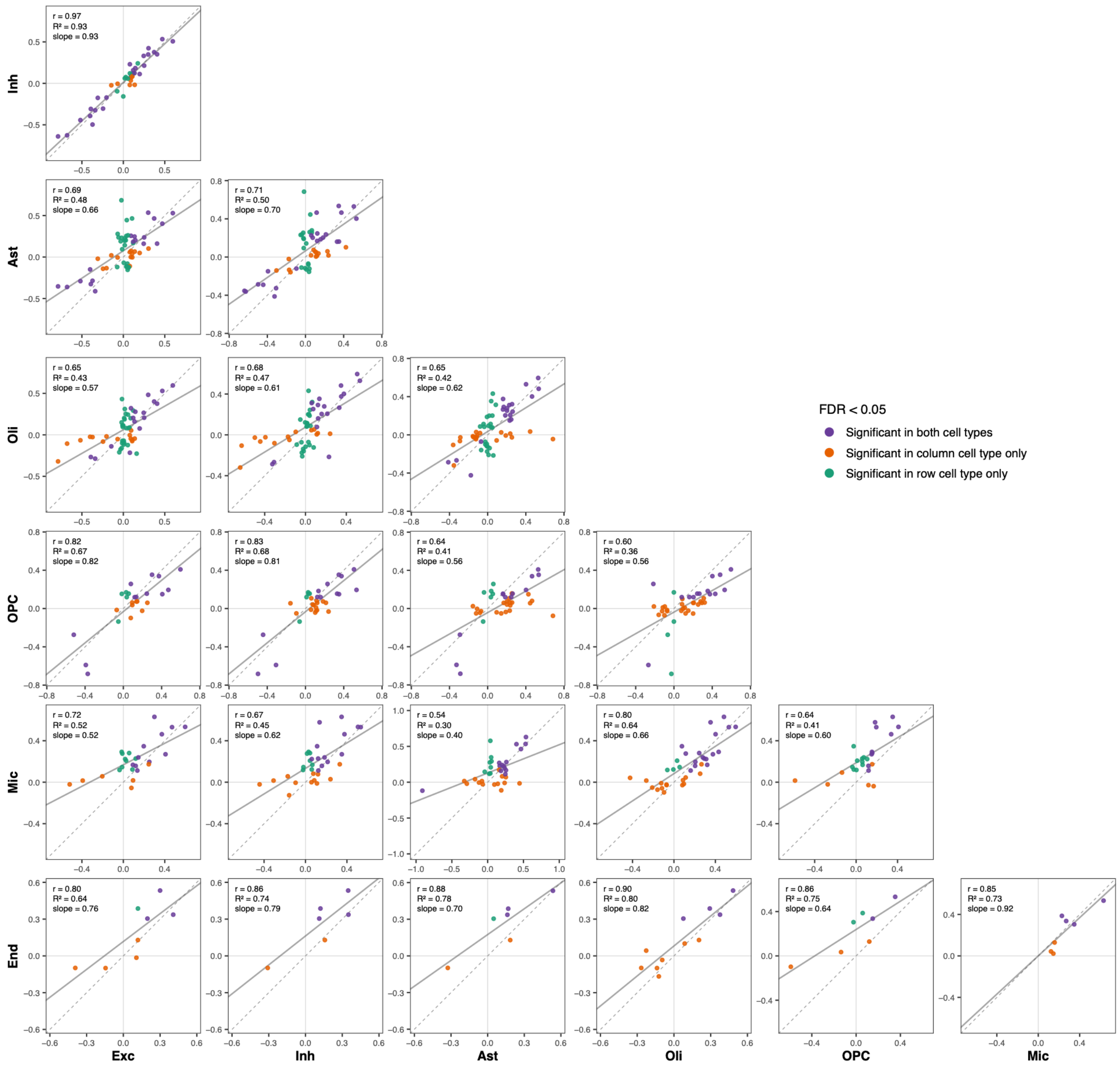

**Supplementary Figure S5. Correlation of sex differences in gene expression across seven cell types.** Each panel compares sex-DEG effect sizes ($\log_2$ fold-change [$\log_2$FC], female vs male; logFC > 0 indicates female-upregulation and logFC < 0 male-upregulation) between two cell types. For each comparison, DEGs significant (FDR < 0.05) in at least one of the two cell types were included. Six genes involved in X-chromosome inactivation (*XIST*, *TSIX*, *JPX*, *FTX*, *RLIM*, *XACT*) were excluded from all comparisons. Abbreviations: Exc, excitatory neurons; Inh, inhibitory neurons; Ast, astrocytes; Oli, oligodendrocytes; OPC, oligodendroglial progenitor cells; Mic, microglia; End, endothelial cells.

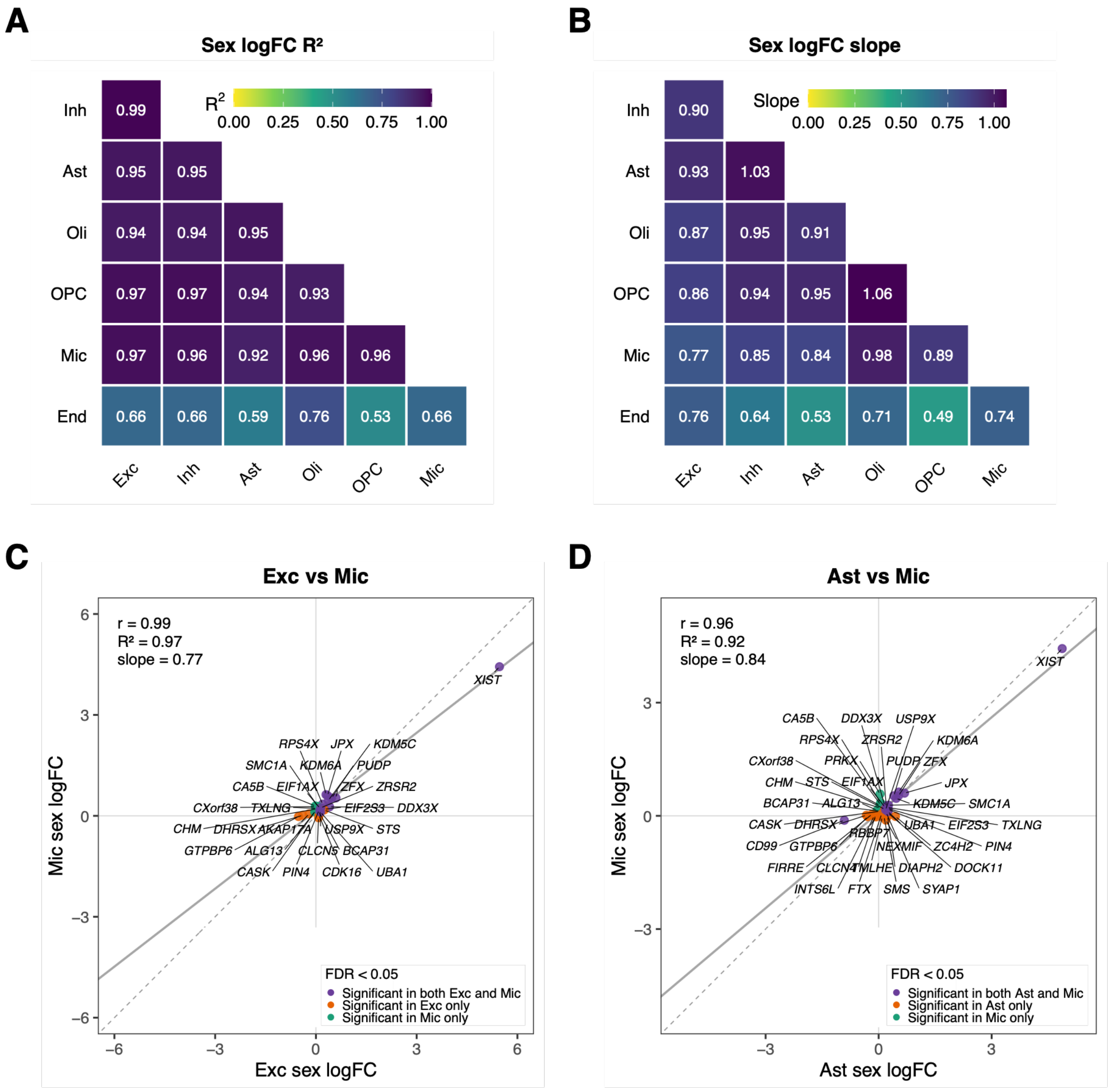

**Supplementary Figure S6. Correlation of sex-DEG effect sizes across cell types without excluding core XCI-regulatory genes.** This analysis repeats **Figure 3J-M** without removing the six genes involved in X-chromosome inactivation (*XIST*, *TSIX*, *JPX*, *FTX*, *RLIM*, *XACT*). **A)** $R^2$ and **B)** slope of the linear fit of sex-DEG $\log_2$ fold-changes ($\log_2$FC) between each pair of cell types, including DEGs significant (FDR < 0.05) in at least one of the two cell types. **C-D)** Scatter plots of sex-DEG $\log_2$FC values for **C)** excitatory neurons versus microglia and **D)** astrocytes versus microglia; genes are colored by whether they are significant in both or only one of the two cell types, and dashed and solid lines indicate the identity line and linear fit, respectively. Abbreviations: Exc, excitatory neurons; Inh, inhibitory neurons; Ast, astrocytes; Oli, oligodendrocytes; OPC, oligodendroglial progenitor cells; Mic, microglia; End, endothelial cells.

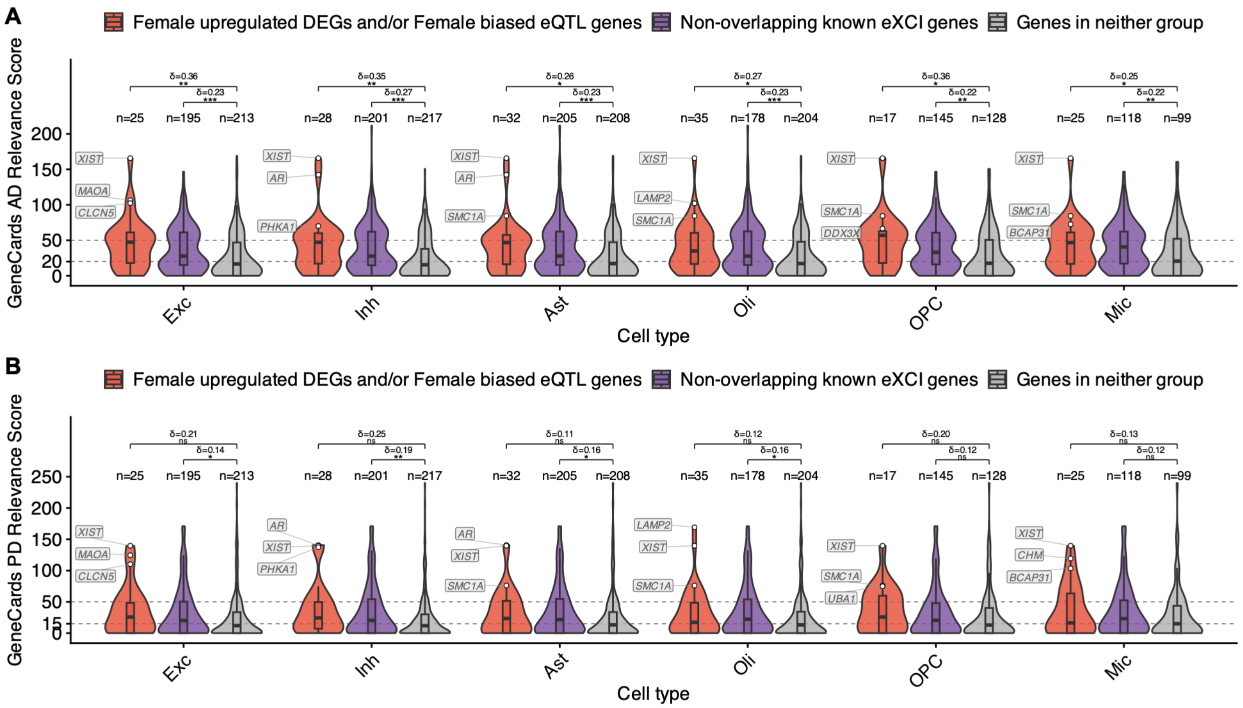

**Supplementary Figure S7. AD and PD relevance of female-biased X-chromosome genes. A** AD relevance scores; **B** PD relevance scores. This analysis repeats the comparison in Figure 4B without removing six genes involved in X-chromosome inactivation (*XIST*, *TSIX*, *JPX*, *FTX*, *RLIM*, *XACT*) from all groups. Violin plots of GeneCards AD/PD relevance scores across three gene groups: female-upregulated DEGs and/or female-biased eQTL genes; non-overlapping previously reported eXCI genes; and genes in neither group, restricted to genes tested in the given cell type. Cliff's delta indicates the effect size of the between-group difference. Within each cell type, the top three female-upregulated DEGs or female-biased eQTL genes are labeled. Asterisks indicate significance levels (*FDR < 0.05; **FDR < 0.01; ***FDR < 0.001; ns, not significant).

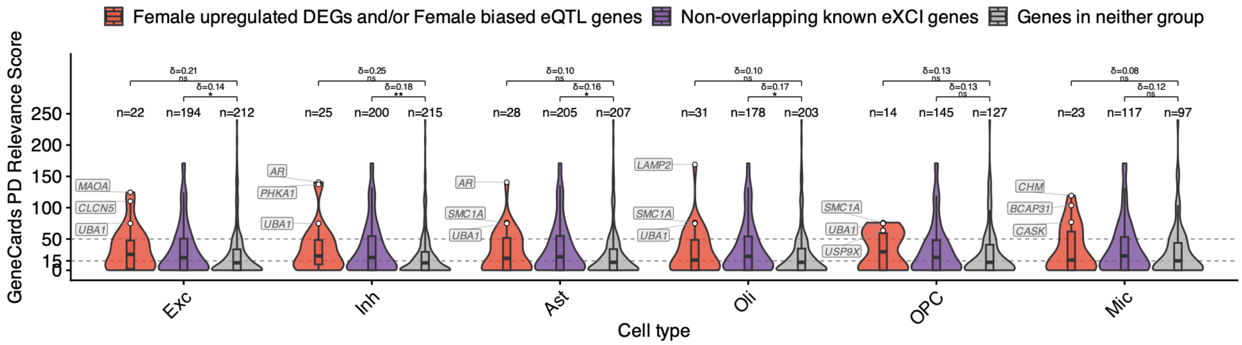

**Supplementary Figure S8. PD relevance of female-biased X-chromosome genes after excluding core XCI-regulatory genes.** This analysis repeats the comparison in Figure S7B after removing six genes involved in X-chromosome inactivation (*XIST*, *TSIX*, *JPX*, *FTX*, *RLIM*, *XACT*) from all groups. Violin plots of GeneCards PD relevance scores across three gene groups: female-upregulated DEGs and/or female-biased eQTL genes; non-overlapping previously reported eXCI genes; and genes in neither group, restricted to genes tested in the given cell type. Cliff's delta indicates the effect size of the between-group difference. Within each cell type, the top three female-upregulated DEGs or female-biased eQTL genes are labeled. Asterisks indicate significance levels (*FDR < 0.05; **FDR < 0.01; ***FDR < 0.001; ns, not significant).

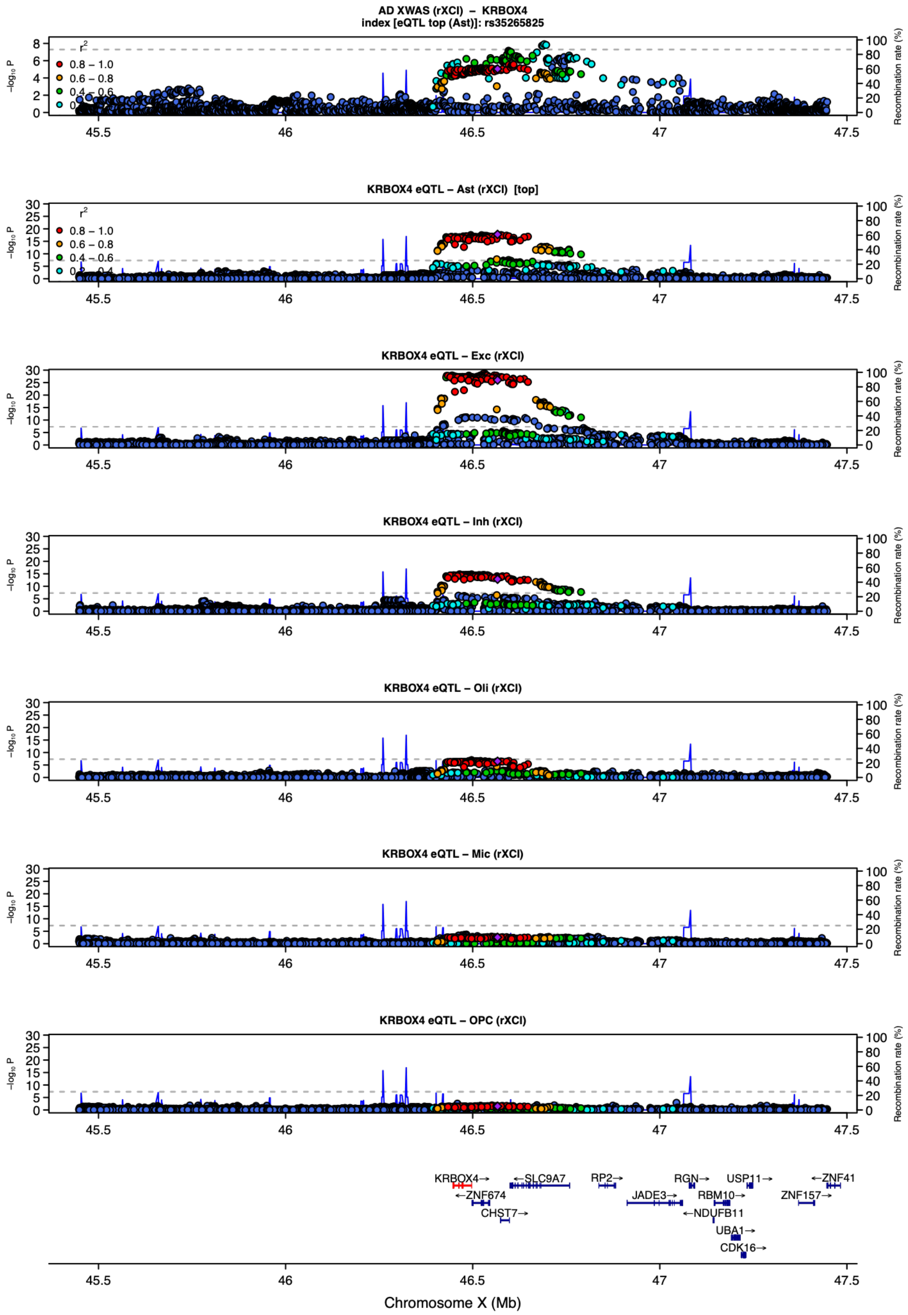

**Supplementary Figure S9. Locus zoom plot for *KRBOX4* (AD).** rXCI XWAS × rXCI eQTL; top cell type, astrocytes. Top panel, AD XWAS signal at the locus; lower panels, cell-type-specific cis-eQTL signals for *KRBOX4*. The plot shows the XWAS-model × eQTL-setting combination yielding the strongest cTWAS association; the top cell type, defined as that with the largest absolute cTWAS Z score, is marked "[top]". Points are colored by LD ($r^2$) with the index variant (diamond), the lead eQTL variant of the top cell type; x axis, chromosome X position (hg38); left y axis, $-\log_{10}(P)$; right y axis, recombination rate. The $-\log_{10}(P)$ axis uses a common scale across all eQTL panels. Cell types: Ast, astrocytes; End, endothelial cells; Exc, excitatory neurons; Inh, inhibitory neurons; Mic, microglia; Oli, oligodendrocytes; OPC, oligodendroglial progenitor cells.

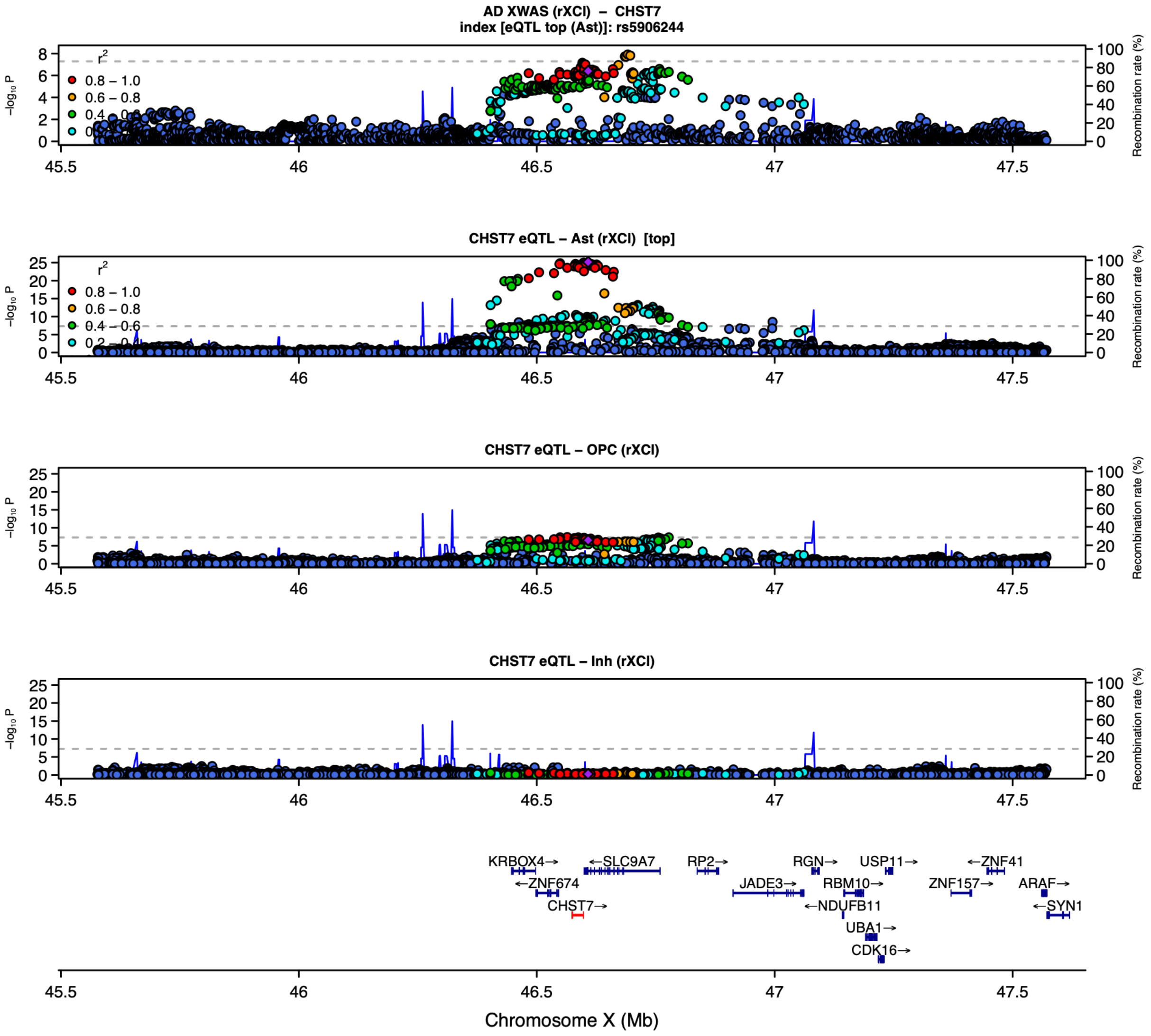

**Supplementary Figure S10. Locus zoom plot for *CHST7* (AD).** rXCI XWAS × rXCI eQTL; top cell type, astrocytes. Panels and coloring as in Supplementary Figure S9.

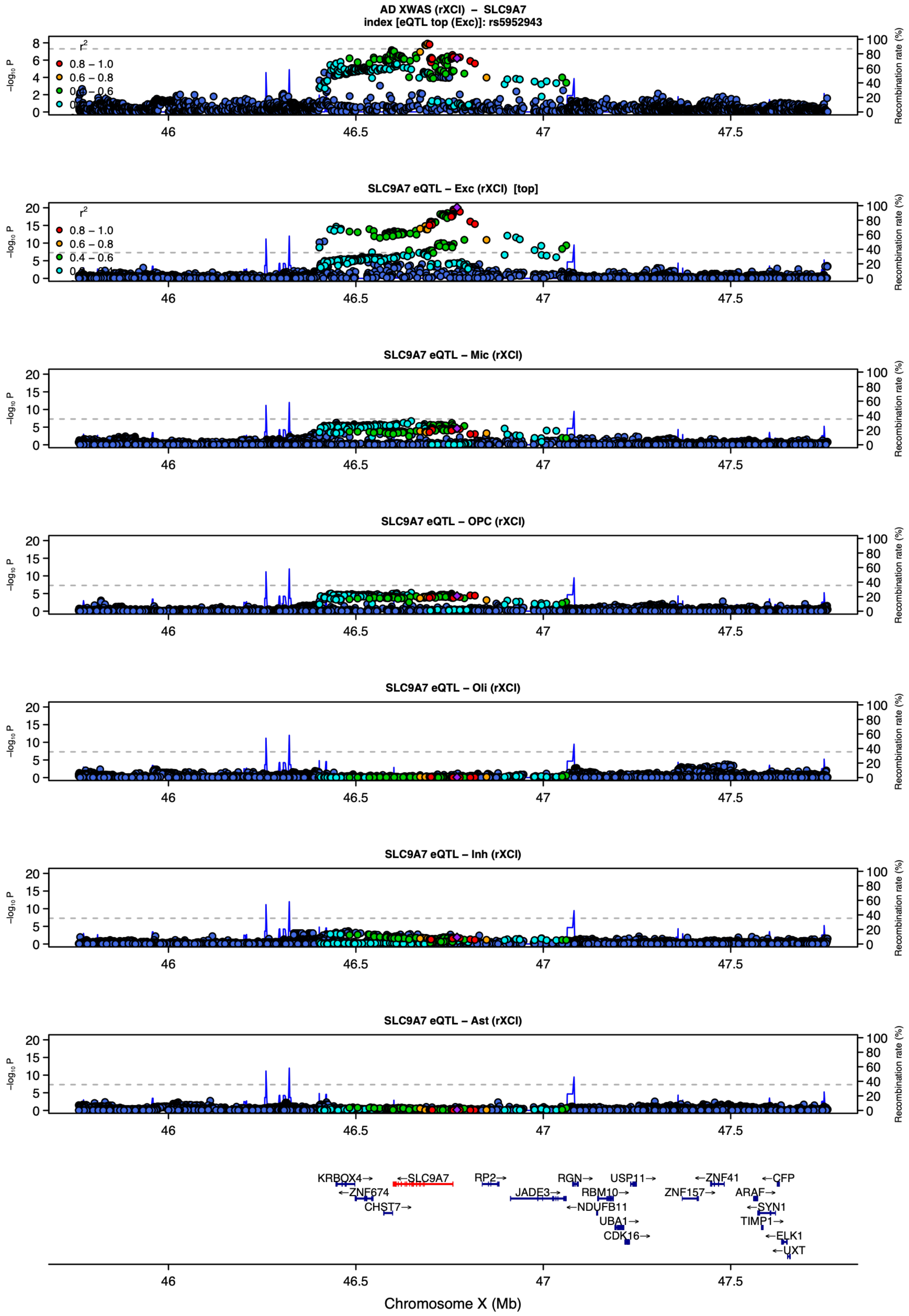

**Supplementary Figure S11. Locus zoom plot for *SLC9A7* (AD).** rXCI XWAS × rXCI eQTL; top cell type, excitatory neurons. Panels and coloring as in Supplementary Figure S9.

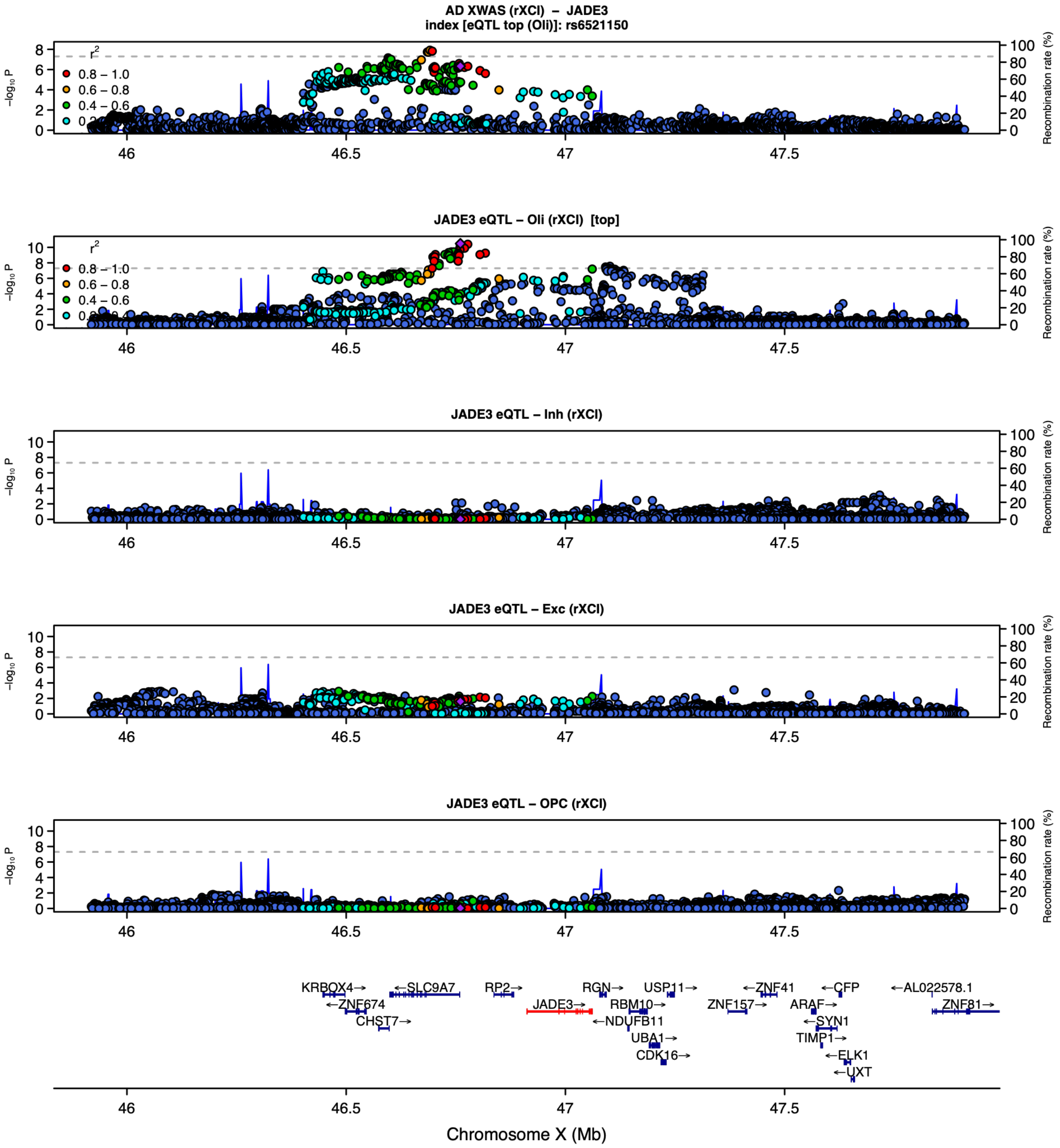

**Supplementary Figure S12. Locus zoom plot for *JADE3* (AD).** rXCI XWAS × rXCI eQTL; top cell type, oligodendrocytes. Panels and coloring as in Supplementary Figure S9.

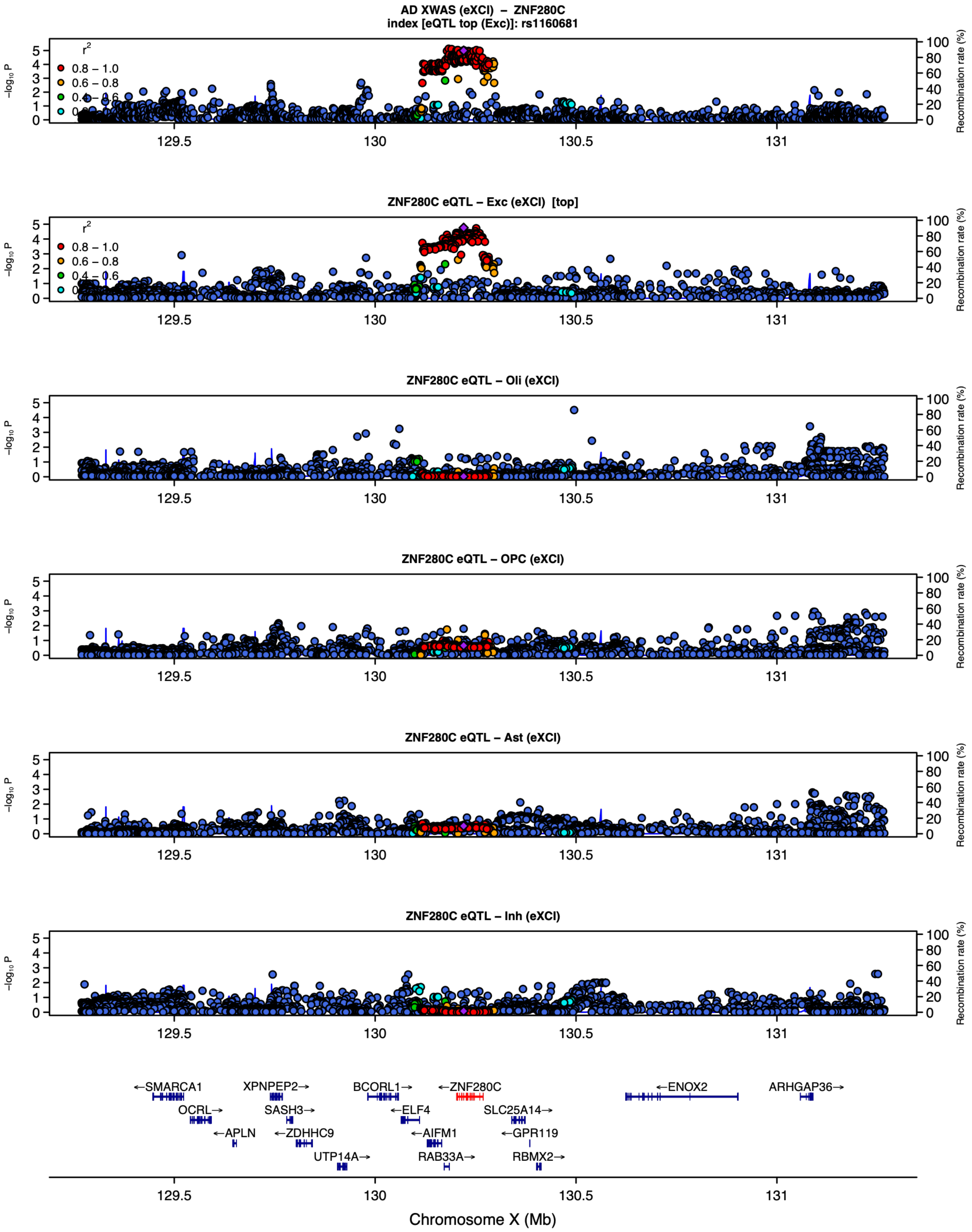

**Supplementary Figure S13. Locus zoom plot for *ZNF280C* (AD).** eXCI XWAS × eXCI eQTL; top cell type, excitatory neurons. Panels and coloring as in Supplementary Figure S9.

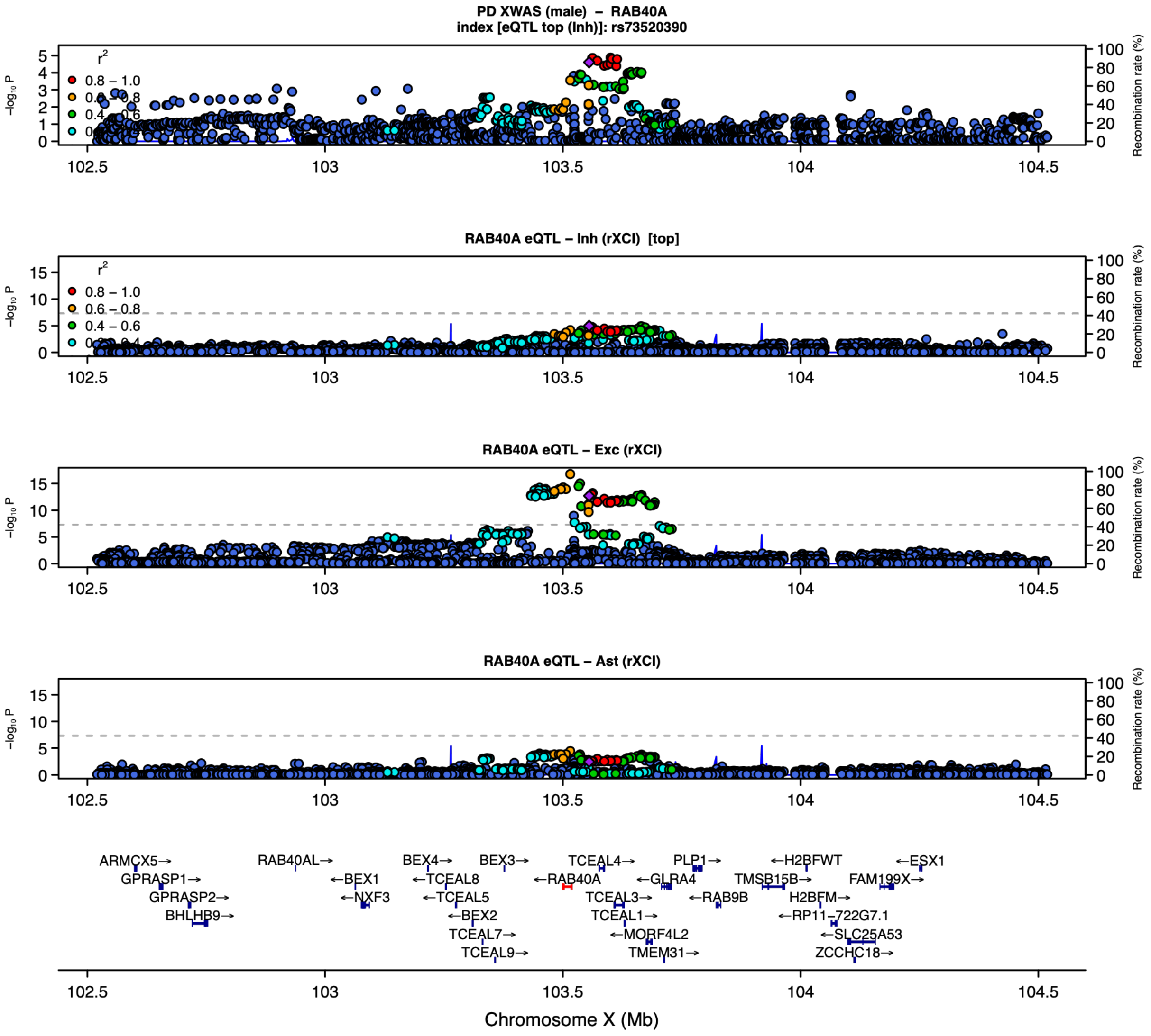

**Supplementary Figure S14. Locus zoom plot for *RAB40A* (PD).** male XWAS × rXCI eQTL; top cell type, inhibitory neurons. Panels and coloring as in Supplementary Figure S9.

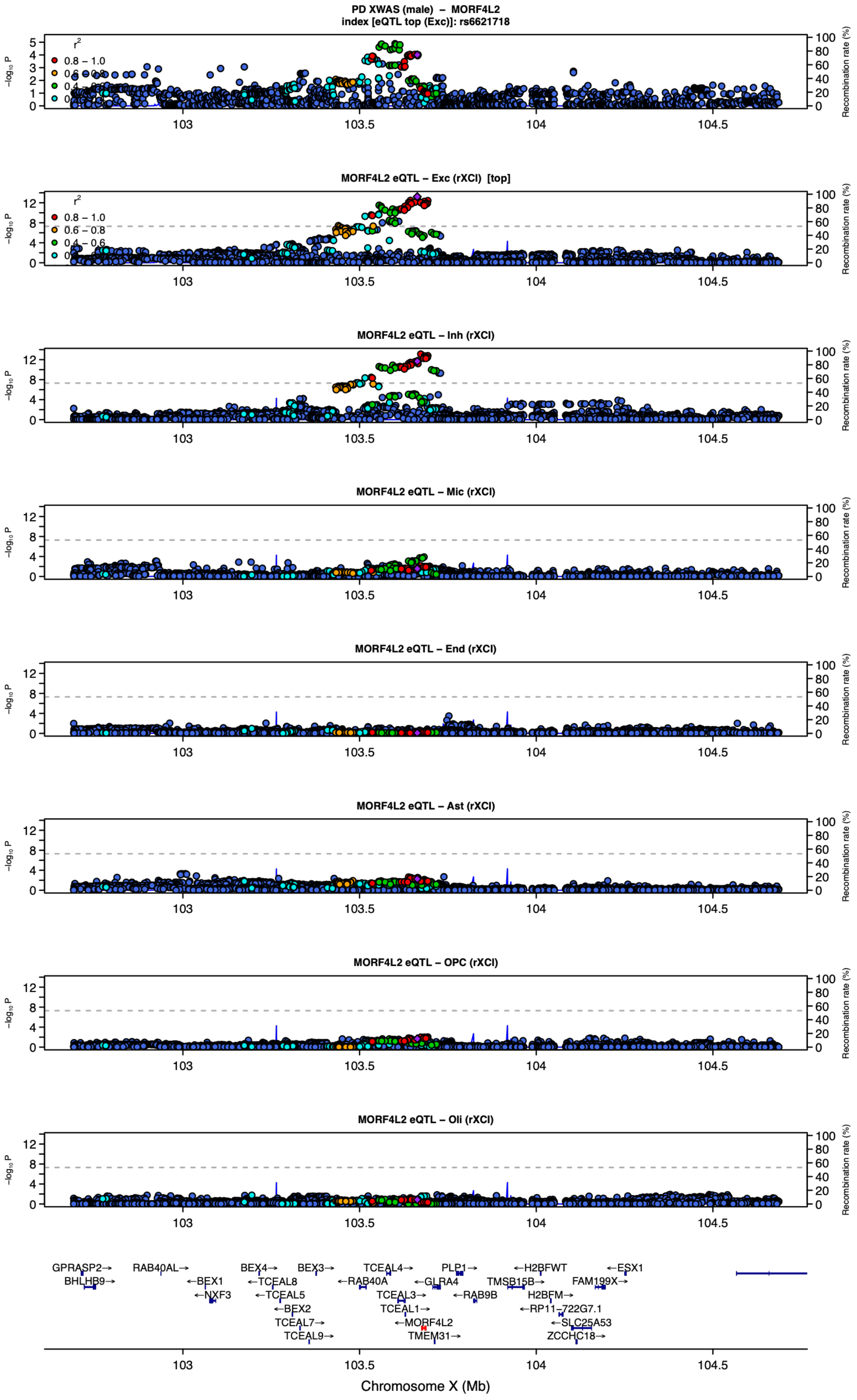

**Supplementary Figure S15. Locus zoom plot for *MORF4L2* (PD).** male XWAS × rXCI eQTL; top cell type, excitatory neurons. Panels and coloring as in Supplementary Figure S9.

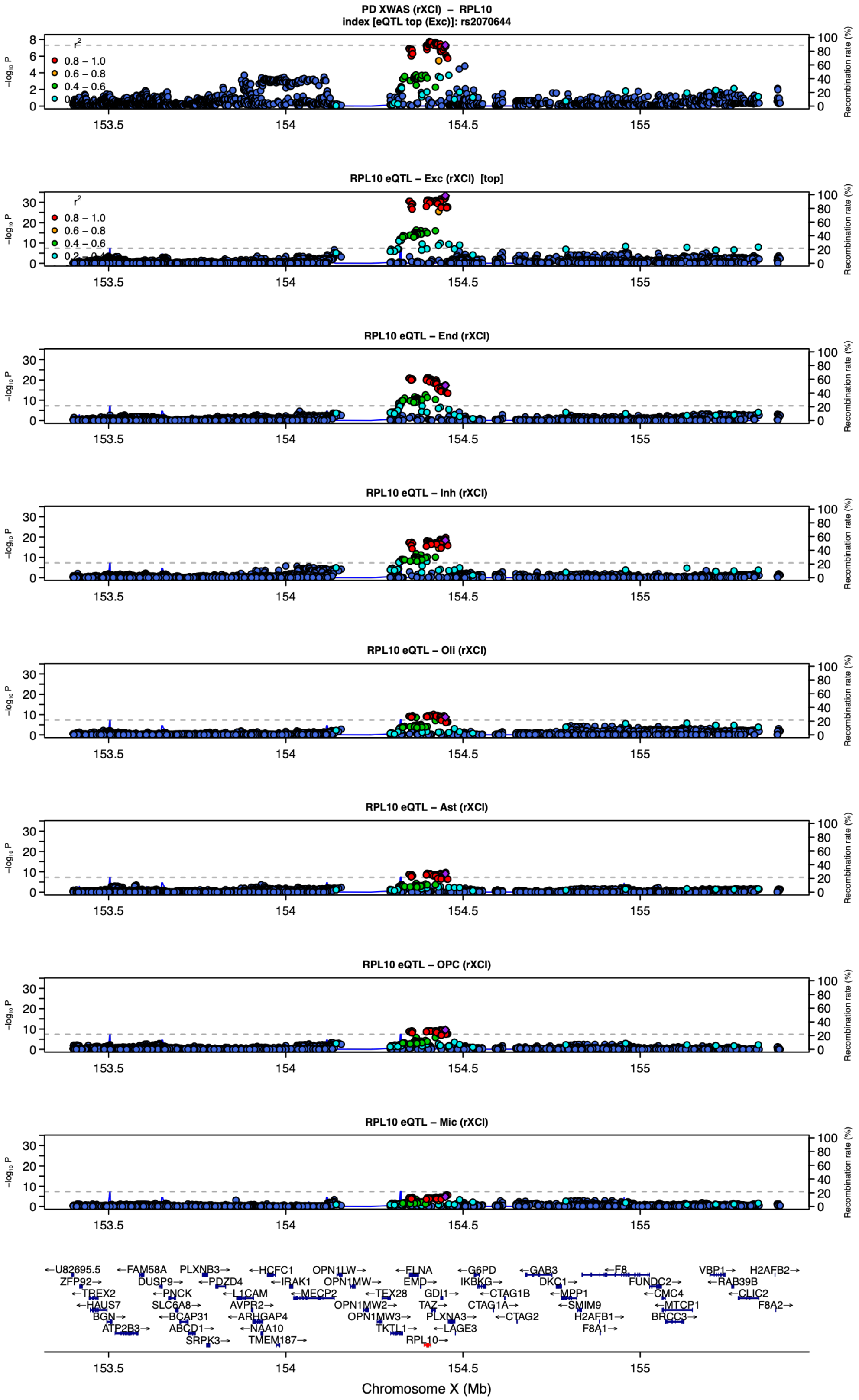

**Supplementary Figure S16. Locus zoom plot for *RPL10* (PD).** rXCI XWAS × rXCI eQTL; top cell type, excitatory neurons. Panels and coloring as in Supplementary Figure S9.

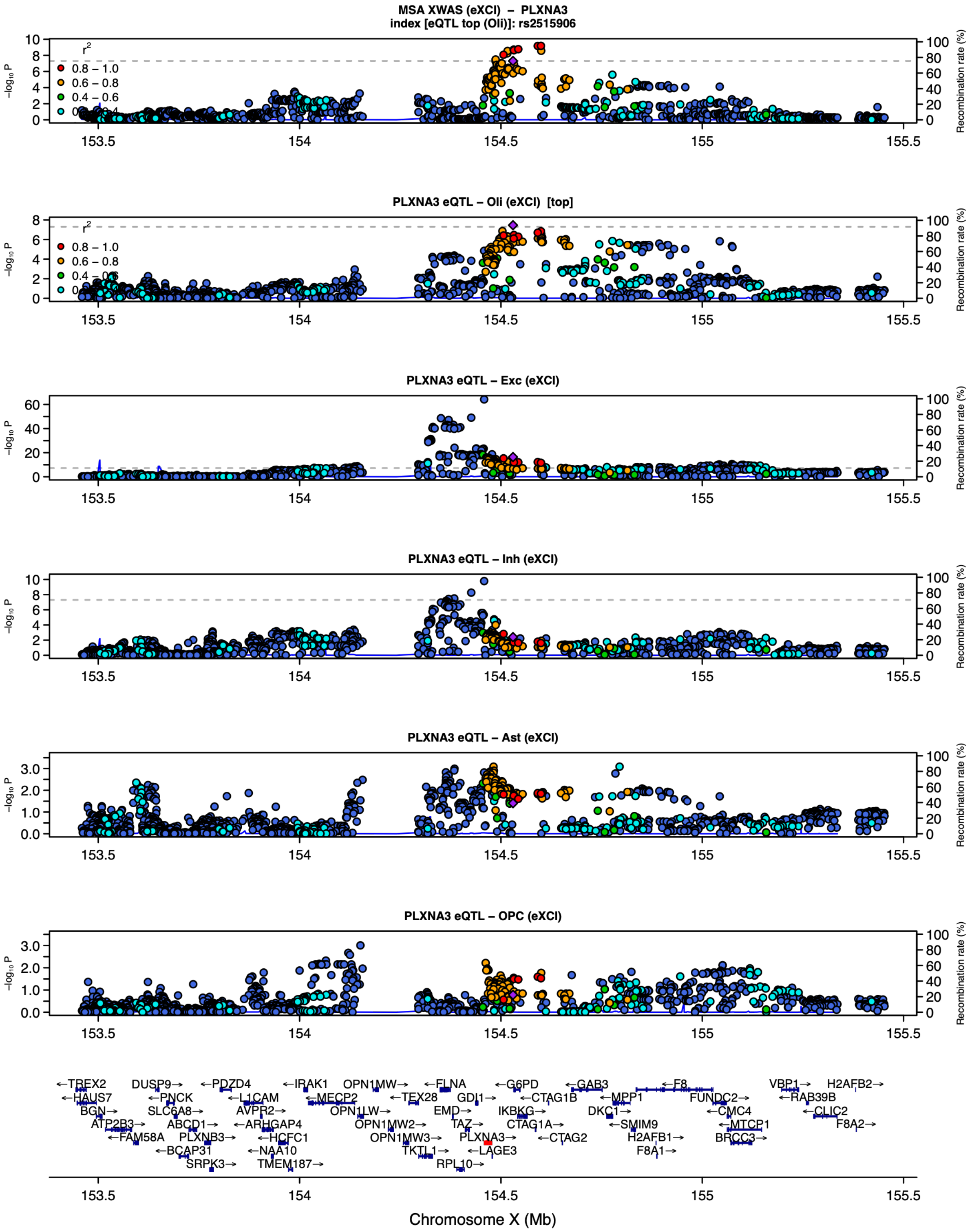

**Supplementary Figure S17. Locus zoom plot for *PLXNA3* (MSA).** eXCI XWAS × eXCI eQTL; top cell type, oligodendrocytes. Panels and coloring as in Supplementary Figure S9, except that the −log10(P) axis is scaled independently for each panel, rather than on the common scale used in the other locus zoom plots, to accommodate the much stronger cis-eQTL of PLXNA3 in excitatory neurons than in oligodendrocytes.

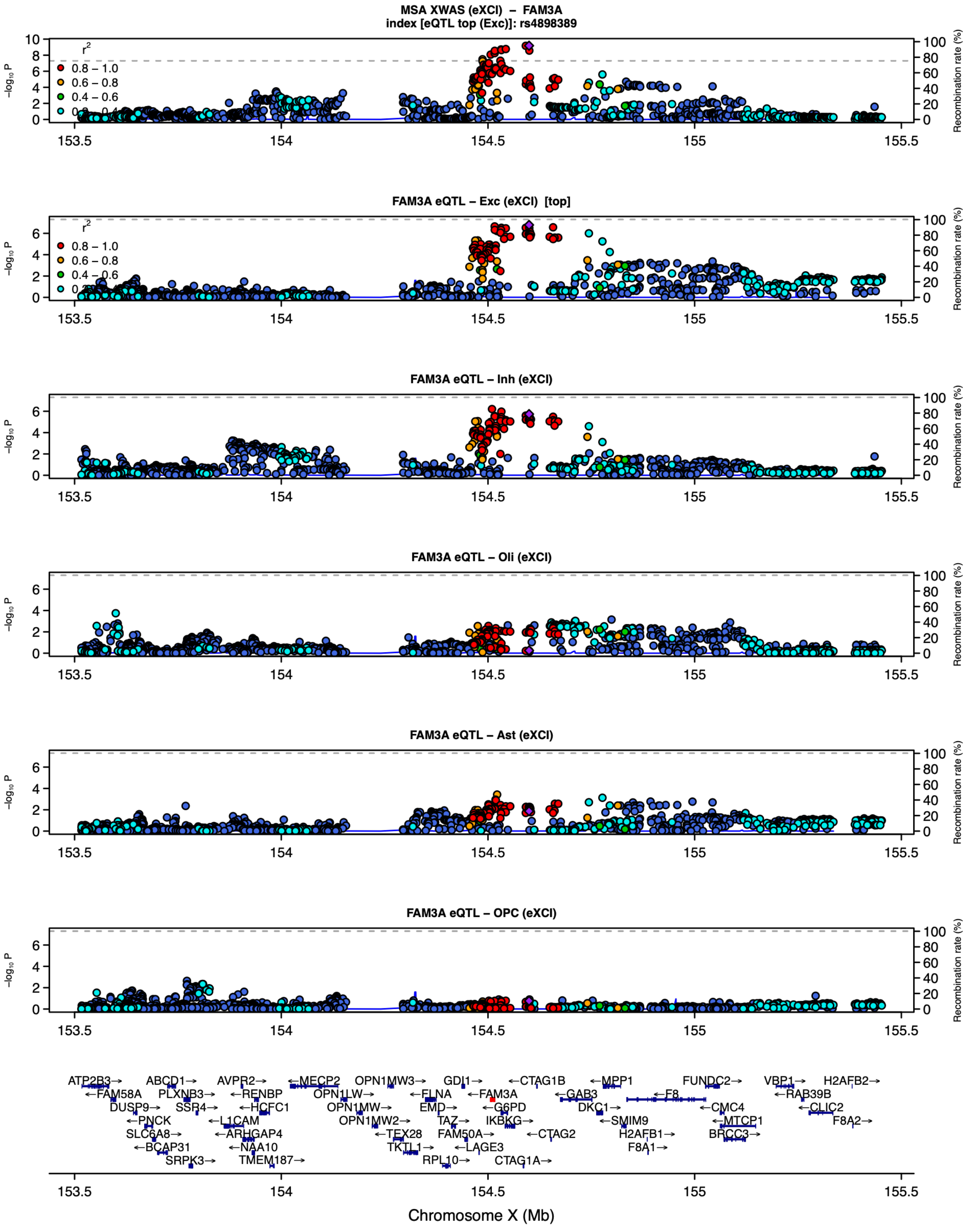

**Supplementary Figure S18. Locus zoom plot for *FAM3A* (MSA).** eXCI XWAS × eXCI eQTL; top cell type, excitatory neurons. Panels and coloring as in Supplementary Figure S9.

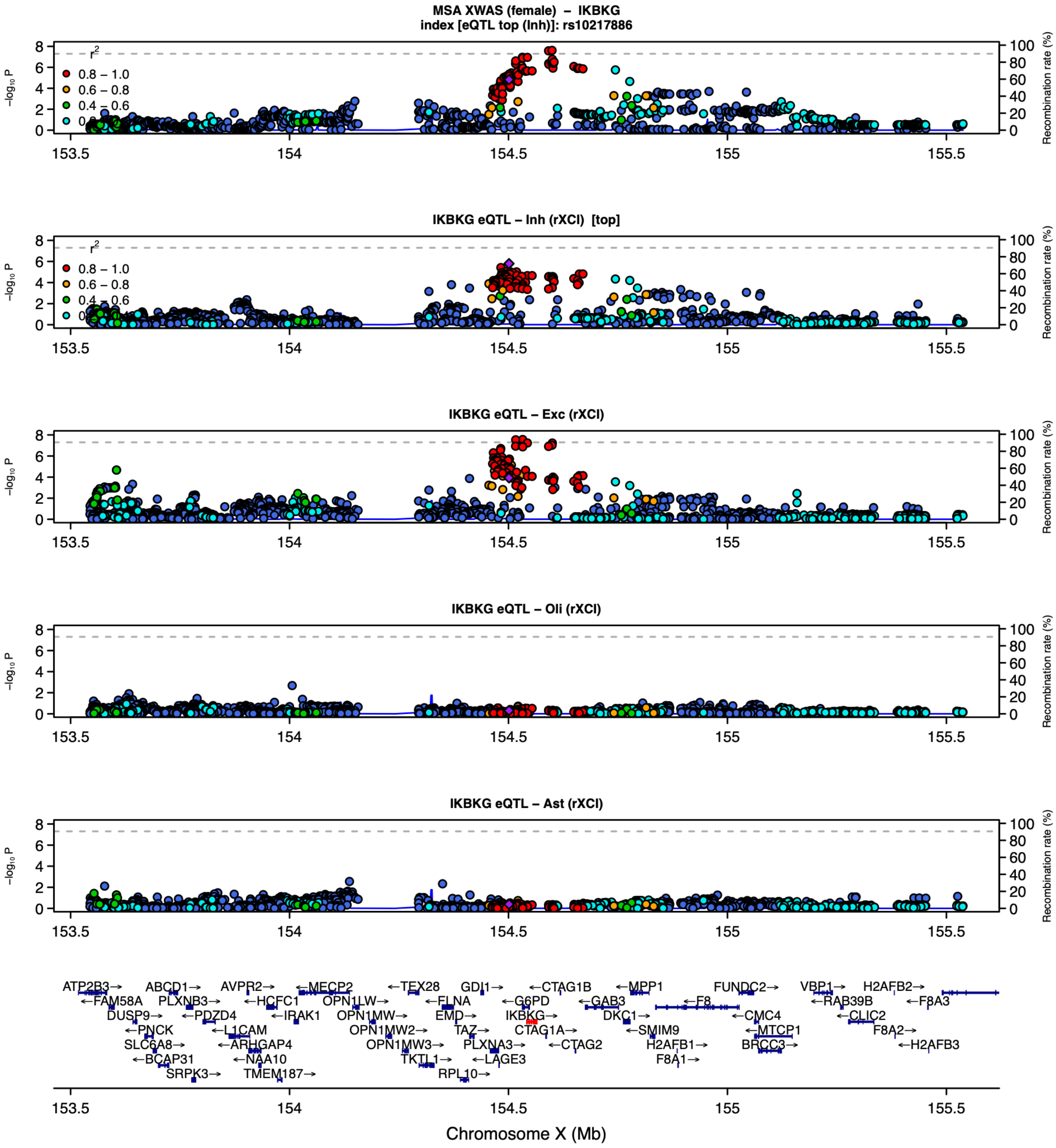

**Supplementary Figure S19. Locus zoom plot for *IKBKG* (MSA).** female XWAS × rXCI eQTL; top cell type, inhibitory neurons. Panels and coloring as in Supplementary Figure S9.
